# Profiles of US Hispanics Unvaccinated for COVID-19

**DOI:** 10.1101/2021.09.17.21263464

**Authors:** Brian J. Piper, Bianca V. Sanchez, Joshua D. Madera, Michael A. Sulzinski

## Abstract

**Background:** The COVID-19 pandemic has disproportionately impacted Hispanics in the US with increased rates of SARS-Cov2 infections, hospitalizations, and deaths. The objective of this report was to characterize the demographics and beliefs of unvaccinated Hispanics to help address their concerns that lead to vaccine hesitancy.

**Methods:** Of 1,011 potential participants from a national online panel, 22.3% (N=225, 51.6% female, age = 40.5) met inclusion criteria of Hispanic adults and not receiving at least one dose of the COVID-19 vaccine. The 30-item survey included items about demographics, political affiliations, sources of news (e.g., Fox vs. CNN), reasons for being unvaccinated, and ratings (0 = strongly disagree, 100 = strongly agree) of 10 controversial statements regarding COVID-19.

**Results:** Over three-fifths (62.6%) identified side effects and safety concerns while almost one-third (30.5%) a lack of efficacy as their top reasons for being unvaccinated. Agreement to “*The developers of the COVID-19 vaccine rushed the development and cut-corners*” was rated highest (63.22) which was significantly (*p* < .001) higher than the other nine statements (e.g., “*The COVID-19 vaccine does not work*”). Many vaccine attitudes differed significantly by political party affiliation and some by gender and news source. Republicans (59.9 ± 4.2) scored higher than Democrats (38.5 ± 4.2, *p* ≤ .001) to “*If I’ve already had COVID-19, I don’t need the vaccine*.”

**Conclusions:** This study identified heterogeneity in COVID-19 vaccine attitudes among Hispanics. Further research is needed to determine if the subgroups identified are differentially receptive to interventions to facilitate reconsideration of prior vaccination decisions.

---

Although the coronavirus disease 2019 (COVID-19) pandemic has affected everyone, the disease burden in the US has disproportionately impacted minorities. A systematic review determined that Hispanic populations had a 1.3 to 7.7 times greater risk for a positive SARS-COV2 (Severe Acute Respiratory Syndrome Coronavirus 2) RNA-PCR relative to non-Hispanic White populations.^1^ Moreover, Hispanic or Latino individuals were 2.7 to 4.4-fold more likely to be hospitalized due COVID-19 and 2.8 fold more likely to die from the disease, relative to non-Hispanic Whites.^1-3^ Further, more years of life were lost due to COVID-19 before age 65 among the Hispanic and non-Hispanic Black populations than Whites, despite the smaller size of these groups.^3^ However, vaccination rates of Hispanics lagged relative to Whites in 34 of the 40 states reporting ethnicity. For example, one-third (33%) of Hispanics versus two-thirds of Whites (64%) had received a COVID-19 vaccine dose in Arizona.^4^

Vaccination decisions are complex and impacted by a variety of cultural, demographic, sociopolitical, religious, and economic factors.^4-13^ A scoping review of ninety-two studies from high-income countries determined that risk for vaccine hesitancy was highest for those of non-White ethnicity, younger-age, females, lower-education, lack of recent history of receiving the influenza vaccination, decreased perceived risk of contracting COVID-19, and not having chronic medical conditions.^10^ Similarly, an online study with a national sample (N=1,878) conducted in 2020 determined that Hispanics, those with children at home, rural residents, and people identifying politically as Republicans were less likely to be vaccinated.^11^ Phone-interviews of Medicare patients completed in the fall of 2020 revealed that those whose primary information source was social media had lower perceptions of COVID-disease severity and lower likelihood of getting a vaccinated.^12^ Four out of five (79.9%) Hispanic/Latino women who were pregnant were unvaccinated for COVID-19.^13^

As the 62.1 million Hispanics constitute the largest minority in the US,^14^ the objectives of this investigation were to extend upon past research^9,11^ to further characterize Hispanics who were unvaccinated for COVID-19 upto August 1, 2021.

## METHODS

### Procedures

Potential participants received an invite from Survey Monkey between July 14 and August 1, 2021. The survey was hosted on this survey firm’s panel which has 2.5 million daily respondents who are compensated ($0.25 – $0.50/survey). Inclusion criteria were identification as Hispanic, adult (age > 18), and a negative response to “Have you received at least one dose of the COVID-19 vaccine, from any maker?”. There were six items about demographics (age, gender, ethnicity, income, education, political affiliation). There were ten statements and misconceptions, e.g. “The COVID-19 vaccine will make me infertile”, selected based on research^15-17^ and the extent of agreement was rated on a 100-point scale. There were six items targeting domains potentially related to vaccine hesitancy including sources of news, future presidential voting preference, vaccination status of others, and perceived ages where the COVID-19 vaccine benefits exceeded the risk. Pilot testing was completed by sending an electronic copy to interested parties. The full-instrument including the five standard Survey Monkey demographic items is available in the a Supplemental Appendix. The CHERRIES was followed for reporting.^18^ Procedures were deemed exempt by the Institutional Review Board of Geisinger in Danville, Pennsylvania.

### Data-analysis

Statistical analysis was completed with Systat, version 13.1. Figures were prepared with GraphPad Prism, version 6.07 with variability depicted as the SEM. When the “prefer not to disclose” option was selected, these participants were removed from the denominator for percentage calculations for that question. Associations between the ten COVID-19 statement ratings were determined with Pearson correlations. Cronbach’s alpha was used for internal consistency and principle component analysis for the ten statements. A *p* < .05 was considered statistically significant although analyses that met more conservative cutoffs (e.g., *p* < .0005) were noted.

## RESULTS

### Participant Characteristics

There were 1,011 potential participants (48.06% female, 42.60% age 18-44 and 15.41% age > 61; 53.80% with an annual income < $50K; 22.87% Pacific, 21.62% South Atlantic, and 19.96% West South Central Census Regions), with 225 (22.26%) meeting the inclusion criteria of not receiving a COVID-19 vaccine.

Half (51.6%) of the sample was female, age = 40.48, SD=14.93, Min=18, Max = 83. Half of respondents selected Mexico (47.91%) followed by Puerto Rico (19.53%), and Cuba (6.05%) to “*I or my family is ethnically from one or more of the following countries*?”. Geographically, the Census regions represented included the South Atlantic (25.25%), Pacific (23.27%), West South Central (18.32%), and Middle Atlantic (11.39%) regions. Half (48.37%) had a personal annual income ≤ $40K. The education of half (49.76%) was high-school or less. The political affiliation was approximately evenly divided between Democratic (31.76%), Republican (31.18%), and Independent (30.59%). The mean response to “*If the election were held today, how likely would you vote for Donald Trump or Joe Biden?”*, with Biden = 0 and Trump = 100, was 47.51 (SD = 38.89). Three-fifths (61.43%) selected “does not apply to me” to “*Within the past month, how often have you attended in person or virtual religious services?*”. Fox (51.46%), traditional broadcasters (ABC, CBS, & NBS = 51.46%), social media (46.20%), CNN (45.03% including CNN en espanol = 11.11%), the local newspaper (15.79%), and Telemundo (15.79%) were each selected as among the top three primary sources of news (Supplementary Figure 1).

### Reasons for Non-Vaccination

Table 1 shows a ranking for the top three reasons for not receiving the COVID-19 vaccine. Over three-fifths endorsed concern about side effects and safety. Three out of ten indicated that they do not believe it will protect them from COVID-19. Over one-quarter did not believe it was necessary because they had a prior COVID-19 diagnosis or suspected one. One out of eleven reported a medical exemption. Religious beliefs were endorsed by one-ninth. Logistical issues like cost, transportation to the vaccination site, obtaining time off work, or difficulty signing up for a vaccination were each selected by less than 8%. Among the eighteen participants that elected to provide an “other” reason, these were varied and included “allergy” or “autoimmune disease” (three responses), “believe it’s a placebo”, “don’t like needles”, “pregnant”, “I have a healthy body”, “I just don’t want to”, or “haven’t had a reason to”.

**Table 1.** Ranking of responses to “What are your top three reasons for not receiving the COVID-19 vaccine?” among US Hispanics.

|  | <u>Percent</u> |
| --- | --- |
| 1. Concerned about side effects | 62.57% |
| 2. Safety concerns about vaccine contents | 62.57% |
| 3. Don't believe it will protect me from COVID-19 | 30.48% |
| 4. Don't believe it is necessary (previously COVID-diagnosed) | 16.58% |
| 5. Most everyone else around me has received the vaccine | 16.04% |
| 6. Religious beliefs | 11.23% |
| 7. Don't believe it is necessary (suspect previous COVID-19) | 10.16% |
| 8. Medical exemption | 9.09% |
| 9. Cost | 7.49% |
| 10. Lack of transportation to vaccination site | 5.88% |
| 11. Difficulty getting time off work | 5.35% |
| 12. Don't know how to sign up for a vaccination | 5.35% |

The response to “*How likely would you be to take the COVID vaccine if it were a pill*?” with options ranging from 0 to 100% were generally low (Mean = 32.01%, SD = 34.41%, Median = 16.50%) but higher for Democrats (44.36%, SD = 34.08) than Republicans (24.37%, SD = 30.33, *t*(103) = 3.17, *p* < .005). Independents (30.89, SD = 35.61) were less likely than Democrats (*t*(103) = 1.98, *p* ≤ .05).

Social contributions to vaccine hesitancy were investigated by asking “*How many of the 30 people you interact with (non-virtually) the most each week (your bubble) have received the COVID vaccine?*” with options from 0 to 100% were intermediate (Mean = 39.23%, SD = 29.46, Median = 42.50). Males (43.24%, SD = 29.13) indicated that more of their interactions were with vaccinated people than females (33.85%, SD = 27.81, *t*(200) = 2.34, *p* < .05). The subsequent question was “*How likely would you be to receive the vaccine if the majority of your bubble received the vaccine?*” produced a modest value (Mean = 30.89%, SD = 32.45, Median = 19.00) with 29.72% of participants selecting 0%. Males (38.82%, SD = 32.63) were higher than females (22.86%, SD = 29.52, *t*(200) = 3.65, *p* < .0005). Democrats (43.04%, SD = 31.52) were elevated relative to Republicans (27.19%, SD = 30.86, *t*(103) = 2.60, *p* < .05) and also Independents (24.39%, SD = 30.83, *t*(103) = 3.07, *p* < .005).

### Age and Subgroup Dependency of Vaccination

Overall, the responses to “*For what ages and groups do the COVID-19 vaccine benefits exceed the risks or side effects?*” were age dependent with less than one-fifth endorsing vaccination for minors (newborns and age 1-4: 17.22%, age 5-11: 15.31%, age 12-17: 16.75%) which then gradually increased (18-29: 24.4%, 30-49: 32.06%, 50-64: 39.71%) with highest values for the elderly (> 65: 49.28%). Vaccination of pregnant woman was favored by slightly over one-fifth (21.05%).

### COVID-19 Beliefs & Misconceptions

Ten controversial COVID statements were ranked on a 0 (strongly disagree) to 100 (strongly agree) scale. Figure 1 shows that the belief that “*The developers of the COVID-19 vaccine rushed the development and cut-corners*” was rated highest (63.22) which was significantly (*p* < .001) higher than the other nine-statements. The only other statement to score greater than 50 (i.e. neutral) was “The COVID-19 vaccine does not work” (51.04) which was rated significantly (*p* < .05) higher than statements in the fourth to tenth rank. The third highest ranking (48.13) was for “If I’ve already had COVID-19, I don’t need the vaccine” which had a significantly (*p* < .05) elevated score relative to statements ranked fifth and below.

**Figure 1.**
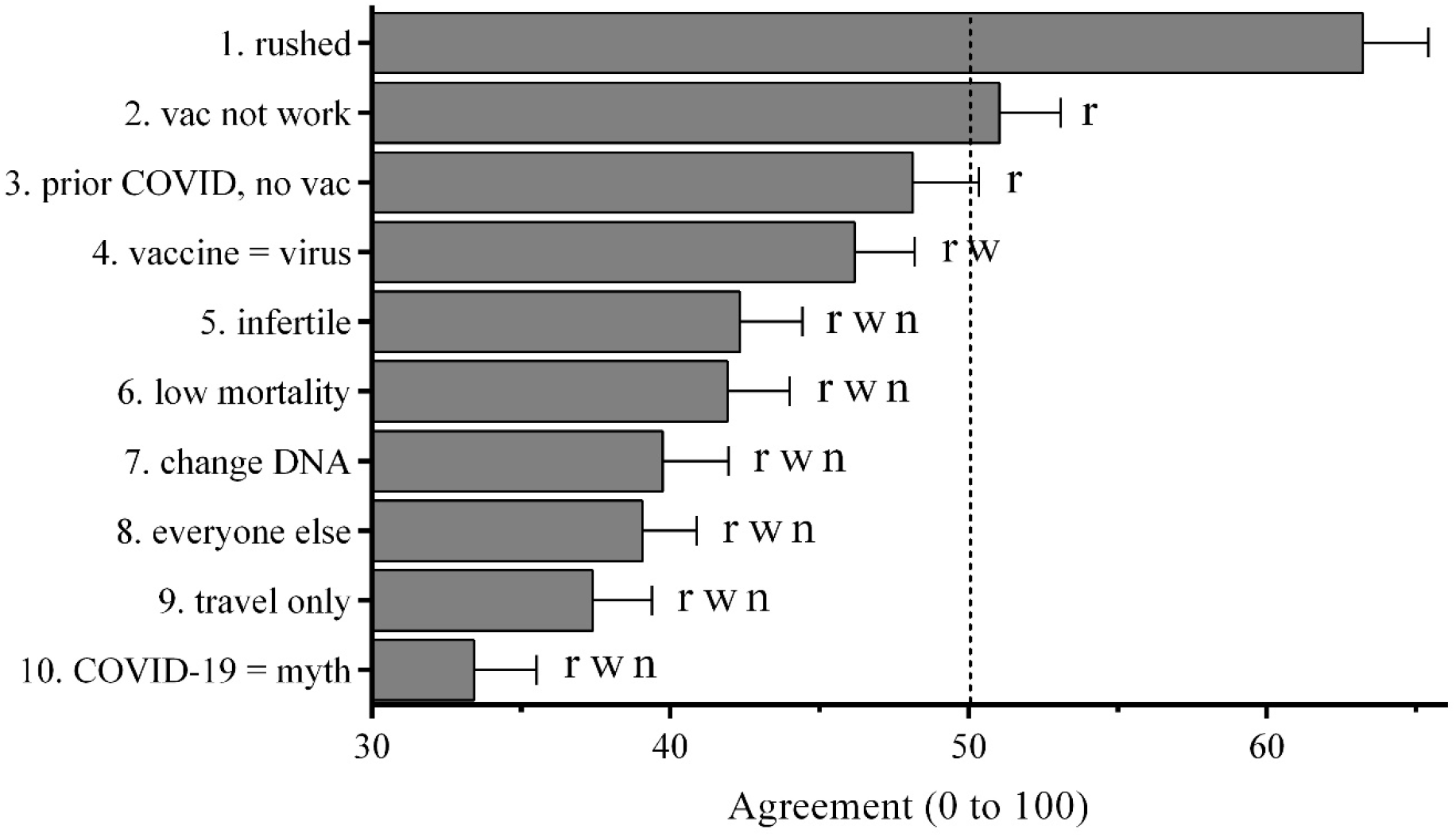
Rating (+SEM) of ten controversial COVID-19 disease and vaccine statements (0 = strongly disagree to 100 = strongly agree) among unvaccinated Hispanics, ranked. Neutral (50) is shown with a vertical dashed line. Statements were: The developers of the COVID-19 vaccine rushed the development and cut corners (1. rushed), The COVID-19 vaccine does not work (2. vac not work), If I’ve already had COVID-19, I don’t need the vaccine (3. prior COVID, no vac), The COVID-19 vaccine is just the virus and will infect you with the disease (4. vaccine = virus), The COVID-19 vaccine will make me infertile (5. infertile), The vaccine isn’t necessary because COVID-19 has a low mortality rate (6. low mortality), The COVID-19 vaccine will change parts of my DNA (7. change DNA), I don’t need the vaccine because everyone else around me has already received it (8. everyone else), I only need the vaccine if I want to travel out of the country (9. travel only), COVID-19 is a myth (10. COVID-19 = myth). ^r^*p* < .001 versus 1. rushed, ^w^*p* < .05 versus 2. vaccine not work, ^n^*p* < .05 versus 3. prior COVID, no vaccine.

Table 2 shows generally moderate (r = 0.3 to 0.6) correlations among these statements with the partial exception of “I only need the vaccine if I want to travel out of the country.” The internal consistency of these ten items was 0.874 which showed only a modest improvement (0.883) with the travel item removed. An exploratory principal component analysis was completed. The first two components accounted for 47.85% and 12.20%, respectively, of the variance. The first component constituted all items except for “travel” which had a high negative loading on the second component (Supplemental Table 1).

**Table 2.** Correlations among ten COVID-19 disease and vaccine statements (0 = strongly disagree to 100 = strongly agree) among unvaccinated Hispanics. Statements were: The developers of the COVID-19 vaccine rushed the development and cut corners (1. rushed), The COVID-19 vaccine does not work (2. not work), If I’ve already had COVID-19, I don’t need the vaccine (3. prior COVID), The COVID-19 vaccine is just the virus and will infect you with the disease (4. vac = virus), The COVID-19 vaccine will make me infertile (5. infertile), The vaccine isn’t necessary because COVID-19 has a low mortality rate (6. low mortality), The COVID-19 vaccine will change parts of my DNA (7. change DNA), I don’t need the vaccine because everyone else around me has already received it (8. everyone else), I only need the vaccine if I want to travel out of the country (9. travel only), COVID-19 is a myth (10. COVID = myth). All correlations were *p* < .001 except ^n^non-significant.

|  | <u>1</u> | <u>2</u> | <u>3</u> | <u>4</u> | <u>5</u> | <u>6</u> | <u>7</u> | <u>8</u> | <u>9</u> | <u>10</u> |
| --- | --- | --- | --- | --- | --- | --- | --- | --- | --- | --- |
| 1. rushed | 1.00 |  |  |  |  |  |  |  |  |  |
| 2. not work | 0.56 | 1.00 |  |  |  |  |  |  |  |  |
| 3. prior COVID | 0.47 | 0.56 | 1.00 |  |  |  |  |  |  |  |
| 4. vaccine = virus | 0.45 | 0.42 | 0.31 | 1.00 |  |  |  |  |  |  |
| 5. infertile | 0.50 | 0.44 | 0.36 | 0.52 | 1.00 |  |  |  |  |  |
| 6. low mortality | 0.38 | 0.57 | 0.50 | 0.42 | 0.44 | 1.00 |  |  |  |  |
| 7. change DNA | 0.44 | 0.54 | 0.55 | 0.46 | 0.54 | 0.53 | 1.00 |  |  |  |
| 8. everyone else | 0.27 | 0.43 | 0.44 | 0.43 | 0.43 | 0.48 | 0.55 | 1.00 |  |  |
| 9. travel only | 0.01<sup>n</sup> | 0.12<sup>n</sup> | 0.25 | 0.23 | 0.15<sup>n</sup> | 0.23 | 0.21<sup>n</sup> | 0.49 | 1.00 |  |
| 10. COVID = myth | 0.23 | 0.50 | 0.37 | 0.47 | 0.36 | 0.55 | 0.53 | 0.45 | 0.28 | 1.00 |

A total score for agreement to these ten controversial COVID statements was created which was 29.4% higher for Republicans than Democrats and also elevated relative to Independents. Table 3 shows that Republicans and Democrats differed significantly on twice as many items (six) as Republicans and Independents (three). There was a significant correlation between total score and likelihood of voting for Donald Trump in the next presidential election (*r*(207) = 0.33, *p* < .0005). However, the total score did not differ by gender or age (not shown). Only those with a graduate or professional education had a mean above 500 (i.e., on the “agree” end of the spectrum for all items, Supplemental Figure 2).

**Table 3.** Ratings of ten COVID-19 disease and vaccine statements (0 = strongly disagree to 100 = strongly agree) among unvaccinated Hispanics, by political party identification. Statements were: The developers of the COVID-19 vaccine rushed the development and cut corners (1. rushed), The COVID-19 vaccine does not work (2. vaccine not work), If I’ve already had COVID-19, I don’t need the vaccine (3. prior COVID), The COVID-19 vaccine is just the virus and will infect you with the disease (4. vac = virus), The COVID-19 vaccine will make me infertile (5. infertile), The vaccine isn’t necessary because COVID-19 has a low mortality rate (6. low mortality), The COVID-19 vaccine will change parts of my DNA (7. change DNA), I don’t need the vaccine because everyone else around me has already received it (8. everyone else), I only need the vaccine if I want to travel out of the country (9. travel only), and COVID-19 is a myth (10. COVID-19 = myth).

|  | Republicans (R) |  | Independents (I) |  | Democrats (D) |  | R vs I | R vs D |
| --- | --- | --- | --- | --- | --- | --- | --- | --- |
|  | <u>Mean</u> | <u>SEM</u> | <u>Mean</u> | <u>SEM</u> | <u>Mean</u> | <u>SEM</u> | <u>p value</u> | <u>p value</u> |
| 1. rushed | 75.0 | 3.8 | 59.9 | 4.6 | 58.0 | 4.6 | .013 | .005 |
| 2. vaccine not work | 60.1 | 3.2 | 50.3 | 4.2 | 48.1 | 4.6 | .084 | .047 |
| 3. prior COVID | 59.9 | 4.2 | 52.4 | 4.6 | 38.5 | 4.3 |  | .001 |
| 4. vaccine = virus | 55.7 | 4.3 | 42.6 | 4.0 | 46.4 | 3.8 | .029 |  |
| 5. infertile | 45.5 | 4.1 | 38.3 | 4.2 | 41.2 | 4.6 |  |  |
| 6. low mortality | 52.2 | 4.2 | 42.3 | 4.2 | 35.6 | 4.3 | .099 | .007 |
| 7. change DNA | 52.8 | 4.2 | 36.8 | 4.6 | 35.9 | 4.5 | .012 | .007 |
| 8. everyone else | 41.9 | 3.2 | 38.6 | 3.5 | 33.0 | 3.9 |  | .080 |
| 9. travel only | 38.2 | 4.0 | 33.3 | 3.9 | 38.9 | 4.4 |  |  |
| 10. COVID-19 = myth | 41.1 | 4.4 | 35.5 | 4.2 | 28.2 | 4.0 |  | .033 |
| Total 1 to 10 | 522.5 | 24.5 | 430.0 | 29.5 | 403.9 | 30.2 | .018 | .003 |

The total score was examined based on news source. Those whose primary source of news was CNN had a lower score relative to Fox, NBC, CNN en Espanol (*p* < .05), and local newspaper (*p* < .01, Supplemental Figure 3). Fox viewers were neutral (52.9, SD = 31.4) while CNN viewers slightly disagreed (37.7, SD = 28.4) regarding “*If I already had COVID-19, I don’t need the vaccine*” (*t*(144) = 2.95, *p* < .005). Fox news (67.4, SD = 29.2) viewers scored higher than CNN (52.9, SD = 31.5) on the “vaccine was rushed” statement (*t*(144) = 2.86, *p* < .005).

## DISCUSSION

This novel report with a national US sample of unvaccinated Hispanics is generally congruent with and extends upon prior COVID-19 vaccine hesitancy research conducted earlier in the pandemic and with less targeted samples.^8,9,11,12,16,17^ Two complementary approaches were used to identify the rationale for not being vaccinated eight months after the first vaccine had received an emergency use authorization. First, when participants were asked to select their top three-reasons, concern about side-effects and safety concerns regarding the vaccine contents were identified by over three-fifths of participants. Side-effects were the primary concern even before a COVID vaccine was available.^8^ A small subset, one out of eleven, endorsed religious beliefs. There was a negative association between an external health locus of control and vaccination intentions^5^ as well as misconceptions about fetal tissue being used in vaccine production^19^ so this reported frequency was lower than anticipated. One out of twelve participants selected “cost” which is curious as the vaccine is freely provided, perhaps revealing an important misconception that could be targeted. Continued educational efforts on how to sign-up for the vaccination or greater use of mobile clinics or increased vaccination availability by primary care providers may be practical strategies to target these small (< 6% each), but important, unvaccinated subgroups. The subset (9.1%) of participants reporting a medical exemption may also warrant further attention as the Centers for Disease Control currently recommends vaccination for everyone > age 12^20CDC^ with no absolute contraindications.

Our second strategy to identify individual differences in vaccination decisions was to ask participants to rate their agreement with ten contentious COVID-19 statements. Interestingly, even among this unvaccinated sample, participants, on average, disagreed that the COVID-19 will make them infertile, will change their DNA, or that the disease is a myth. The statement that was most strongly endorsed was that “*The developers of the COVID-19 vaccine rushed the development and cut corners*.” These quantitative findings are congruent with a large (N = 754) qualitative report from Arkansas.^16^ The second highest rated statement “*The COVID-19 vaccine does not work*.” The continued emergence of new variants makes the earlier randomized controlled trials^21,22^ less helpful for efficacy information but these well-powered datasets are still valuable to inform short-term safety. Some hospitals make publicly available the pronounced over-representation of the unvaccinated among those that were hospitalized^23,24^ which may also combat this misperception.

It is difficult to understate the degree that political ideology has come to overlap with COVID-19 beliefs.^8,9^ Republicans more strongly endorsed three of the ten COVID statements including that COVID will change my DNA and that “*The COVID-19 vaccine is just the virus and will infect you with the disease*.” relative to those who identified as Independents. Republicans and Democrats differed on six items and on the total score for all ten statements. While the strength of attitudes differed by political party, it is important to recognize that all three political affiliations (Republican, Democrat, and Independents) were equally represented among the unvaccinated. Similarly, as reported previously,^8,25^ whether these participants obtained their news from more liberal (e.g. CNN) or more conservative (e.g. Fox) sources differentiated COVID-19 attitudes. Importantly, there is some evidence that vaccination disparities by race/ethnicity have narrowed while disparities by political affiliation have widened.^26^Although it is unfortunate that this pressing medical and public health issue is subsumed within the US culture wars for many, these findings and others^9,11^ indicate that unique messages may continue to need to be differentially targeted to these sub-groups.

Vaccination decisions are due to a variety of sociological and psychological factors including race/ethnicity, political beliefs, rural/urban residence, economic considerations, and the intersection of these characteristics.^11,27^ Hispanics unvaccinated for COVID-19 are non-homogenous and exist on a continuum that includes those that are hesitant (e.g. “wait and see”) or facing logistical barriers (e.g. time off work, transportation) to those that whose views are entrenched and may require substantial education, or employment requirements, to change their behavior. There are broad tools like mandates to get vaccinated as a requirement for employment, education, or travel, and more subtle nudges employed by behavioral economists^28-30^. While recognizing that the relationship between attitudes and behaviors is complex,^31^ utilization of positive, targeted^32^, and culturally responsive messaging on COVID-19 vaccines and using vaccinated Hispanic health-care workers as vaccine ambassadors targeting the themes identified here should be evaluated in controlled research.

Some caveats and future directions are noteworthy. First, this investigation relied on self-reported data from a national sample of one-thousand with two-hundred which met the unvaccinated inclusion criteria, recruited online. Future investigations should also target Hispanics whose primary language is not English. Second, we were initially surprised that religious factors did not rank more prominently as a reason for not being vaccinated. The low attendance at virtual or in-person religious events may reflect either the magnitude of COVID-19 induced disruption of these events or that the sample was atypical on this variable. Third, as is true for any point in time survey, these data reflect the interval (Summer, 2021) of data collection which was before the vaccines received full Food and Drug administration approval. Much clinical, epidemiological, and basic science COVID-19 information is rapidly changing^15,20-22^ which will inform survey items on future attitudinal studies.

In conclusion, the stakes are high for understanding, and overcoming, the multifaceted nuances of vaccine hesitancy among Hispanics and others.^4^ We are cautiously optimistic that this report, and future quantitative and qualitative ones, can empirically inform strategies to most efficiently target a decreasing subset of the US population that is unvaccinated against COVID-19.

## Supporting information

Supplemental survey

Raw_data_deidentified

## Data Availability

A dataset is uploaded.

## Availability of data and material

The raw data and survey were included as supplemental materials submitted 9/11/21 to MedRxiv.

## Ethics approval

The Geisinger IRB approved this study as exempt.

## Abbreviations

(COVID-19): Coronavirus disease 2019
(SARS-COV2): Severe Acute Respiratory Syndrome Coronavirus 2

## Acknowledgements

Thanks to Maureen Murtha, MS, Elizabeth Kuchinski, MPH, and Iris Johnston for technical support and Joseph B. Fraiman, MD and Swathi Gotham, MD for discussions on survey design.

---

**Supplemental Figure 1.**
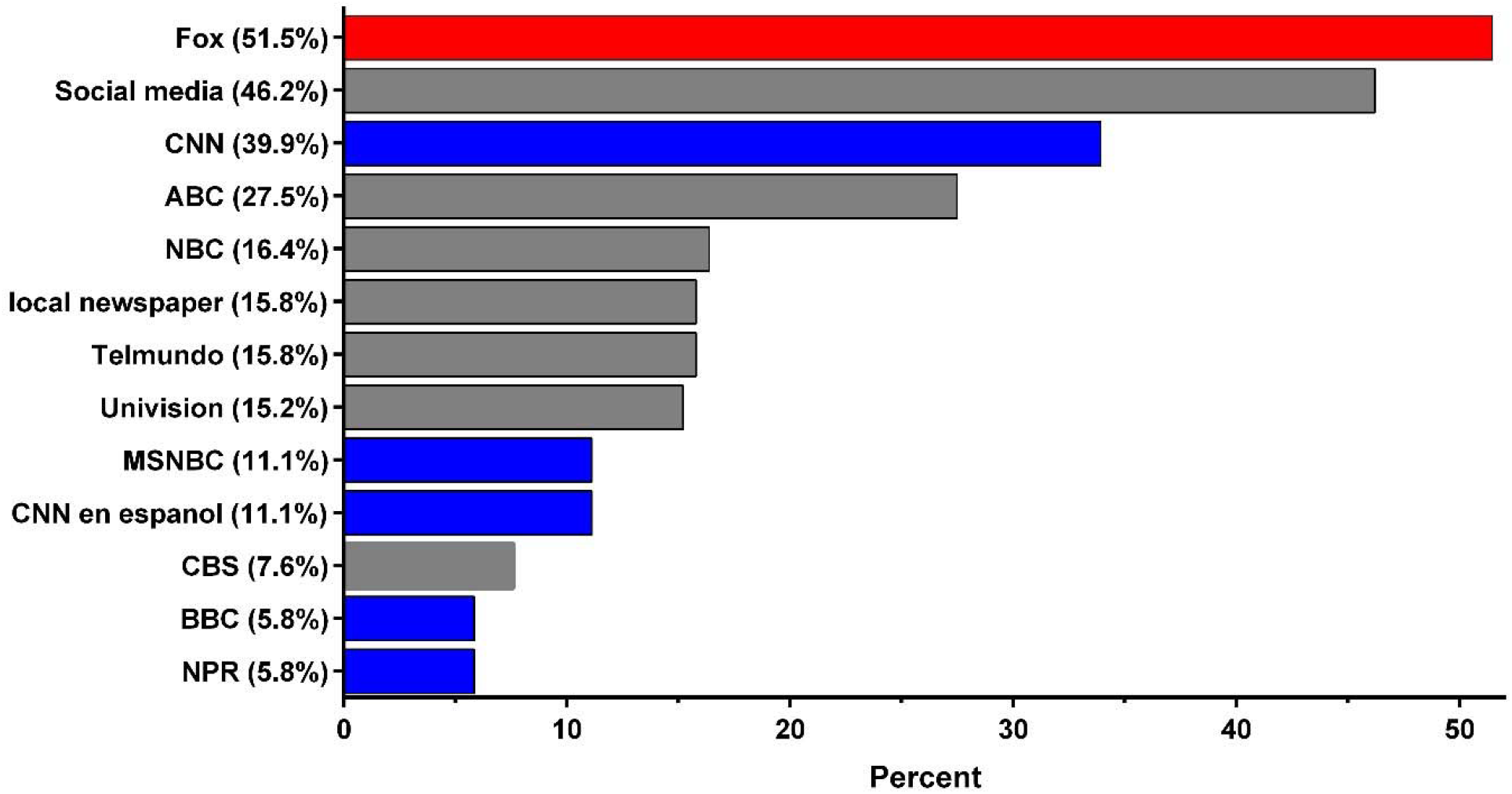
Percent responses to “Which of the following are your top 3 primary sources of news?”, ranked, among COVID-19 unvaccinated Hispanic U.S. respondents (N = 171, N = 37 “prefer not to say” not included). Conservative sources are in **red** and liberal sources in **blue**

**Supplemental Figure 2.**
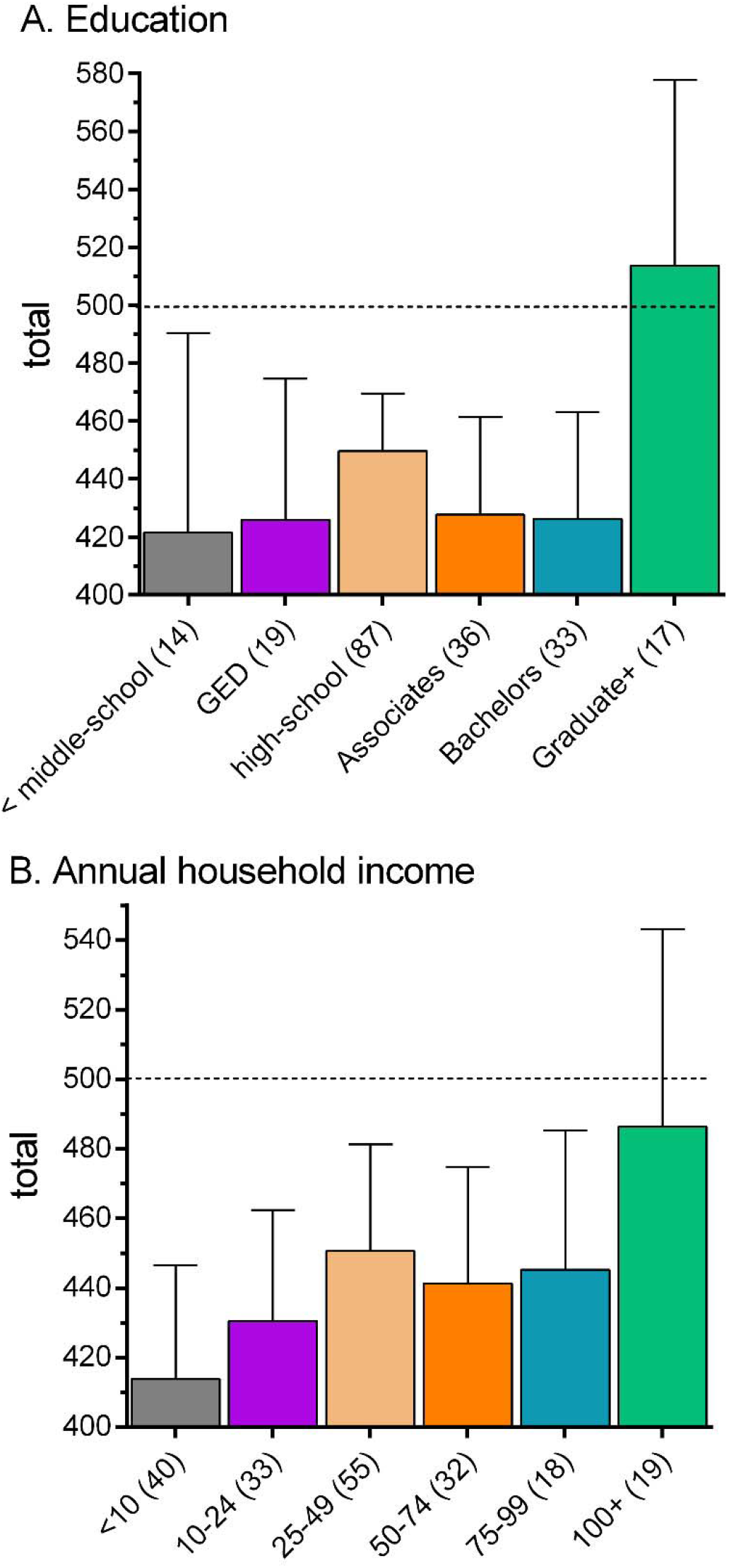
Total score (+SEM) of rating for ten controversial COVID-19 disease and vaccine statements (0 = strongly disagree to 100 = strongly agree) among unvaccinated Hispanics by education (A) and income (B). A total rating of “neutral” on all items is indicated by the horizontal dashed line. The N per group is in parentheses.

**Supplemental Figure 3.**
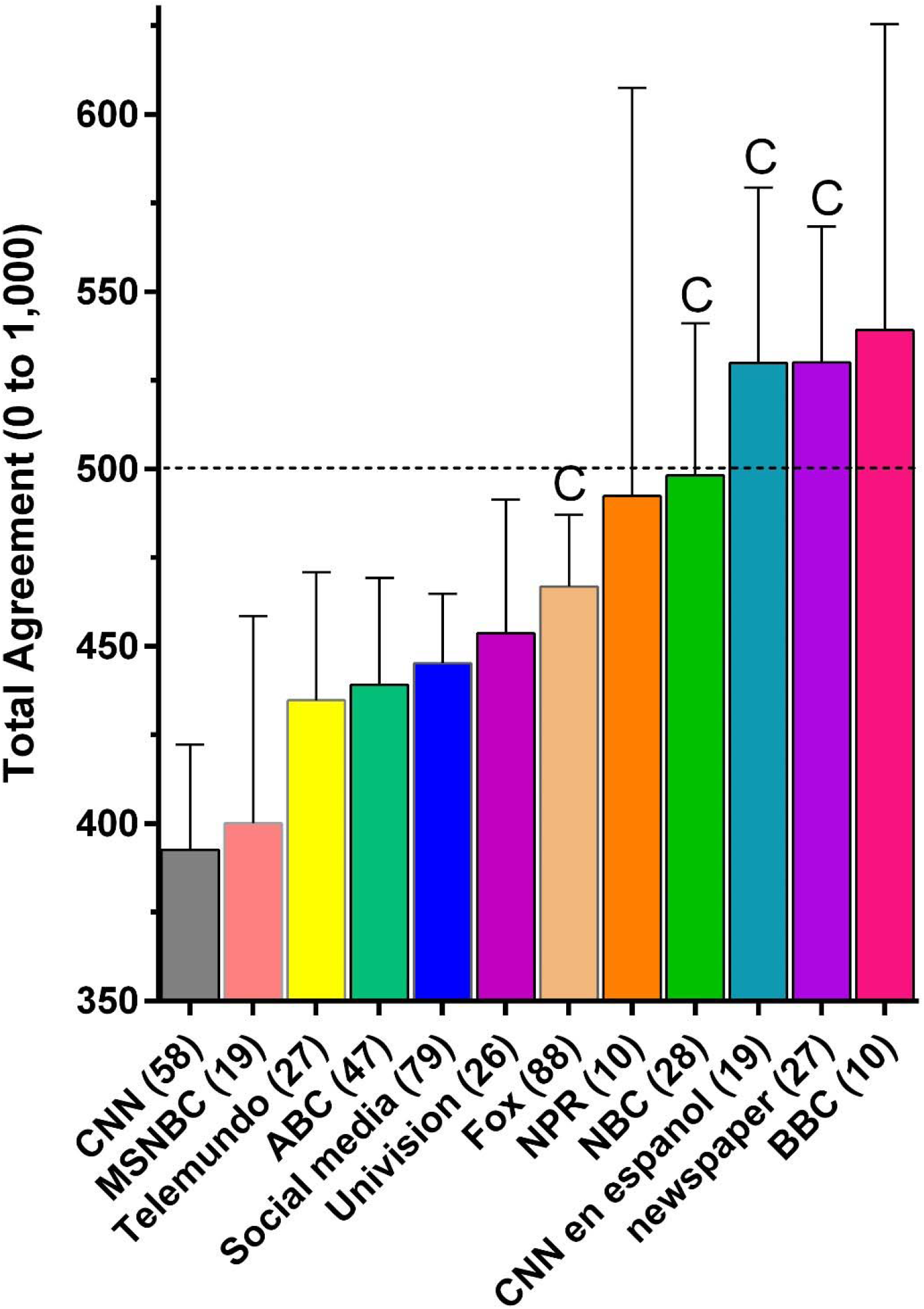
Total score (+SEM) of ratings of ten controversial COVID-19 disease and vaccine statements (0 = strongly disagree to 100 = strongly agree) among unvaccinated Hispanics by top three sources of news. A total rating of “neutral” on all items is indicated by the horizontal dashed line. The N per group is in parentheses. ^C^*p* < .05 versus CNN using https://www.graphpad.com/quickcalcs/ttest2/

**Supplemental Table 1.** Principle component analysis factor loadings for ten controversial COVID-19 disease and vaccine statements (0 = strongly disagree to 100 = strongly agree) among unvaccinated Hispanics. Statements were: The developers of the COVID-19 vaccine rushed the development and cut corners (1. rushed), The COVID-19 vaccine does not work (2. vaccine not work), If I’ve already had COVID-19, I don’t need the vaccine (3. prior COVID), The COVID-19 vaccine is just the virus and will infect you with the disease (4. vaccine = virus), The COVID-19 vaccine will make me infertile (5. infertile), The vaccine isn’t necessary because COVID-19 has a low mortality rate (6. low mortality), The COVID-19 vaccine will change parts of my DNA (7. change DNA), I don’t need the vaccine because everyone else around me has already received it (8. everyone else), I only need the vaccine if I want to travel out of the country (9. travel only), and COVID-19 is a myth (10. COVID-19 = myth). Factor loadings < 0.30 are not shown.

|  | <u>Component 1</u> | <u>Component 2</u> |
| --- | --- | --- |
| 1. rushed | 0.637 | 0.529 |
| 2. vaccine not work | 0.768 |  |
| 3. prior COVID | 0.708 |  |
| 4. vaccine = virus | 0.684 |  |
| 5. infertile | 0.698 |  |
| 6. low mortality | 0.755 |  |
| 7. change DNA | 0.795 |  |
| 8. everyone else | 0.714 | -0.419 |
| 9. travel only | 0.375 | -0.780 |
| 10. COVID-19 = myth | 0.694 |  |

## References

1. Mackey K, Ayers CK, Kondo KK, Sha S, Advani SM, Young S, et al. Racial and ethnic disparities in COVID-19-related infections, hospitalizations, and deaths. Annals Intern Med 2021; 174(3):362–373. doi:10.7326/M20-6306.

2. Centers for Disease Control, COVID-NET. Age-adjusted laboratory-confirmed COVID-19 associated hospitalization rates by race/ethnicity-COVID-NET, March 1, 2020-August 21, 2021. Accessed 9/7/2021 at: https://www.cdc.gov/coronavirus/2019-ncov/images/community/health-equity/COVID-19_assoc-hospital-rates-ethnicity_2021-08-30.jpg

3. Bassett MT, Chen JT, Krieger N. Variation in racial/ethnic disparities in COVID-19 mortality by age in the United States: A cross-sectional study. PLoS Medicine 2020; 17(10):e1003402. doi: 10.1371/journal.pmed.1003402.

4. Ndugga N, Hill L, Artiga S. Latest data on COVID-19 vaccinations by race/ethnicity. Kaiser Family Foundation, Published August 18, 2021. Accessed 9/8/2021 at: https://www.kff.org/coronavirus-covid-19/issue-brief/latest-data-on-covid-19-vaccinations-race-ethnicity/

5. Olagoke AA, Olagoke OO, Hughes AM. Intention to vaccinate against the novel 2019 coronavirus disease: The role of health locus of control and religiousity. J Relig Health 2021; 60(1):65–80. https://doi.org/10.1007/s10943-020-01090-9.

6. Hegarty S. ‘The gospel truth?’ Covid-19 vaccines and the danger of religious misinformation. BBC News. Accessed March 24, 2021 at: https://www.bbc.com/news/av/health-56416683

7. PRRI, IFYC. Religious identities and the race against the virus. Accessed 8/28/2021 at: https://www.prri.org/research/religious-vaccines-covid-vaccination/

8. Ruiz JB, Bell RA. Predictors of intention to vaccinate against COVID-19: Results of a nationwide survey. Vaccine 2021; 1080–6. doi: 10.1016/j.vaccine.2021.01.010.

9. Khubchandani J, Sharma S, Price JH, Wiblishauser MJ, Sharma M, Webb FJ. COVID-19 vaccination hesitancy in the United States: A rapid national assessment. J Community Health 2021; 46(2):270–277. doi: 10.1007/s10900-020-00958-x.

10. Aw J, Benjamin Seng JJ,Ying Seah SS, Low LL. COVID-19 Vaccine hesitancy—A scoping review of literature in high-income countries. Vaccines 2021; 9:900. https://doi.org/10.3390/vaccines9080900.

11. Khubchandani J, Macias Y. COVID-19 vaccination hesitancy in Hispanics and African-Americans: A review and recommendations for practice. Brain Behav Immun Health. 2021;15:100277. doi: 10.1016/j.bbih.2021.100277.

12. Park S, Massey PM, Stimpson JP. Primary source of information about COVID-19 as a determinant of perception of COVID-19 severity and vaccine uptake. J Gen Intern Med 2021; 1–8. DOI: 10.1007/s11606-021-07080-1.

13. Vaccine Safety Datalink. Percent of pregnant people aged 18-49 years receiving at least one dose of a COVID-19 vaccine during pregnancy overall, by race/ethnicity, and date reported to CDC - Vaccine Safety Datalink*, United States. Accessed September 3, 2021 at: https://covid.cdc.gov/covid-data-tracker/?CDC_AA_refVal=https://www.cdc.gov%2Fcoronavirus%2F2019-ncov%2Fcases-updates%2Fspecial-populations%2Fpregnancy-data-on-covid-19.html#vaccinations-pregnant-women

14. US Census Bureau. 2020 Census statistics highlight local population changes and nation’s racial and ethnic diversity. Accessed September 9, 2021 at: https://www.census.gov/newsroom/press-releases/2021/population-changes-nations-diversity.html

15. Crotty S. Hybrid immunity. Science 2021; 372(6549):1392–1393. DOI: 10.1126/science.abj2258.

16. Moore R, Willis DE, Shah SK, Purvis RS, Shields X, McElfish PA. “The risk seems too high”: Thoughts and feelings about COVID-19 vaccination. Int J Environ Res Public Health 2021;18(16):8690. doi: 10.3390/ijerph18168690.

17. Kricorian K, Civen R, Equils. COVID-19 vaccine hesitancy: Misinformation and perceptions of vaccine safety. Hum Vac Immunother 2021-in press; 1–8. doi: 10.1080/21645515.2021.1950504.

18. Eysenbach, G. Improving the quality of Web surveys: The Checklist for Reporting Results of Internet E-Surveys (CHER-RIES). J. Med. Internet Res. 2004;6:e34.

19. Oregon Health Authority. Vaccine facts for Catholic faith communities. Accessed 9/7/21 at: https://sharedsystems.dhsoha.state.or.us/DHSForms/Served/le3592e.pdf

20. CDC. Myocarditis and pericarditis considerations. Accessed 9/8/2021 at: https://www.cdc.gov/vaccines/covid-19/clinical-considerations/myocarditis.html

21. Polack FP, Thomas SJ, Kitchin N, Absalon J, Gurtman A, Lockart S. Safety and efficacy of the BNT162b2 mRNA Covid-19 vaccine. New Eng J Med 2020; 383: 383(27):2603–2615. doi: 10.1056/NEJMoa2034577.

22. Baden LR, El Sahly B, Essink B, Kotloff K, Frey S, Novak R., et al. Efficacy and safety of the mRNA-1273 SARS-CoV-2 vaccine. New Engl J Med 2021; 384(5):403–416. doi: 10.1056/NEJMoa2035389.

23. Burkhalter E. Unvaccinated Alabamians make up majority of people hospitalized with COVID-19. WZDX September 6, 2021. Accessed September 9, 2021 at: https://www.rocketcitynow.com/article/news/local/alabama-icu-shortage-covid-19-coronavirus-vaccine-unvaccinated/525-355418aa-113b-4cc9-80a9-751498831243

24. Johnson N. Over 95 percent of Covid patients at my hospital share one thing in common: They’re unvaccinated. Mens Health September 7, 2021. Accessed September 9, 2021 at: https://www.menshealth.com/health/a37500848/covid-unvaccinated-hospital-doctor-essay/

25. Gollust SE, Vogel RI, Rothman A, Yzer M, Fowler EF, Nagler RH. Americans’ perceptions of disparities in COVID-19 mortality: Results from a nationally-representative survey. Prevent Med 2020; 143:1–9.

26. Szilagyi PG, Thomas K, Shah MD, Vizueta N, Cui Y, Vangala S, Kapteyn A. Likelihood of COVID-19 vaccination by subgroups across the US: Post-election trends and disparities. Hum Vaccin Immunother. 2021;1–6. doi: 10.1080/21645515.2021.1929695.

27. Jackson N. Why some white evangelical Republicans are so opposed to the COVID-19 vaccine. Fivethirtyeight 2021; 8/26/21. Accessed 8/28/21 at: https://fivethirtyeight.com/features/why-some-white-evangelical-republicans-are-so-opposed-to-the-covid-19-vaccine/

28. Shermohammed M, Goren A, Lanyado A, Yesharim R, Wolk DM, Doyle J, et al. Informing patients that they are at high risk for serious complications of viral infection increases vaccination rates. MedRxiv 2021; https://doi.org/10.1101/2021.02.20.21252015.

29. Santos HC, Gore A, Chabris CF, Meyer MN. Effect of targeted behavioral science messages on COVID-19 vaccination registration among employees of a large health system: A randomized trial. JAMA Network Open 2021;4(7):e2118702. doi:10.1001/jamanetworkopen.2021.18702.

30. Dai H, Saccardo S, Han MA, Roh L, Raja N, Vangala S, et al. Behavioural nudges increase COVID-19 vaccinations. Nature. 2021; in press. doi: 10.1038/s41586-021-03843-2.

31. Ajzen I, Fishbein M. The prediction of behavior from attitudinal and normative variable. J Exp Soc Psychol. 1970; 6(4):466–87.

32. Pink SL, Chu J, Druckman JN, Rand DG, Willer R. Elite party cues increase vaccination intentions among Republicans. Proc Natl Acad Sci U S A. 2021;118(32):e2106559118. doi: 10.1073/pnas.2106559118.

