## Supplemental survey for "Profiles of US Hispanics Unvaccinated for COVID-19"

### Online COVID-19 Questionnaire



RULES

+ FILTER + COMPARE + SHOW

#### Your feedback is important to us

Thank you! We read every response. Privacy notice

Do the analyze visualizations and tools help you gather data insights from your survey?

- Yes
- No

Continue

RESPONDENTS: 1,011 of 1,011

ADD TO DASHBOARD SAVE AS

QUESTION SUMMARIES INSIGHTS AND DATA TRENDS INDIVIDUAL RESPONSES

All Pages

Page 1

**Q1** Customize Save as

##### Have you received at least one dose of the COVID-19 vaccine, from any maker?

Answered: 1,011 Skipped: 0

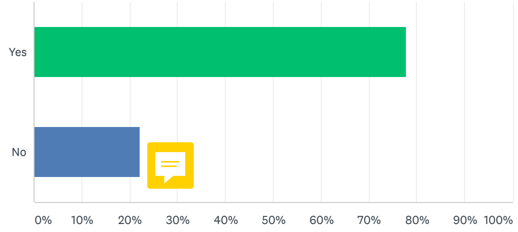


| ANSWER CHOICES | RESPONSES | |
|---|---|---|
| Yes | 77.74% | 786 |
| No | 22.26% | 225 |
| TOTAL | | 1,011 |

Integrate SurveyMonkey with tools you already use, like **Power BI**, **Tableau**, and **Salesforce**, to automate workflows, create deeper insights, and get more value out of your workday.

Get a Demo

Page 2

**Q2** Customize Save as

##### 1. What is your age in years?

Answered: 215 Skipped: 796

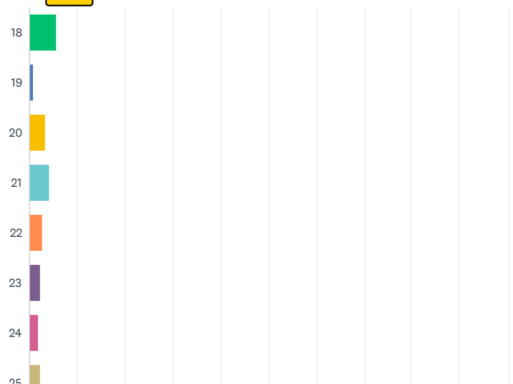

#### Your feedback is important to us

Thank you! We read every response. Privacy notice

Do the analyze visualizations and tools help you gather data insights from your survey?

Yes

No

Continue


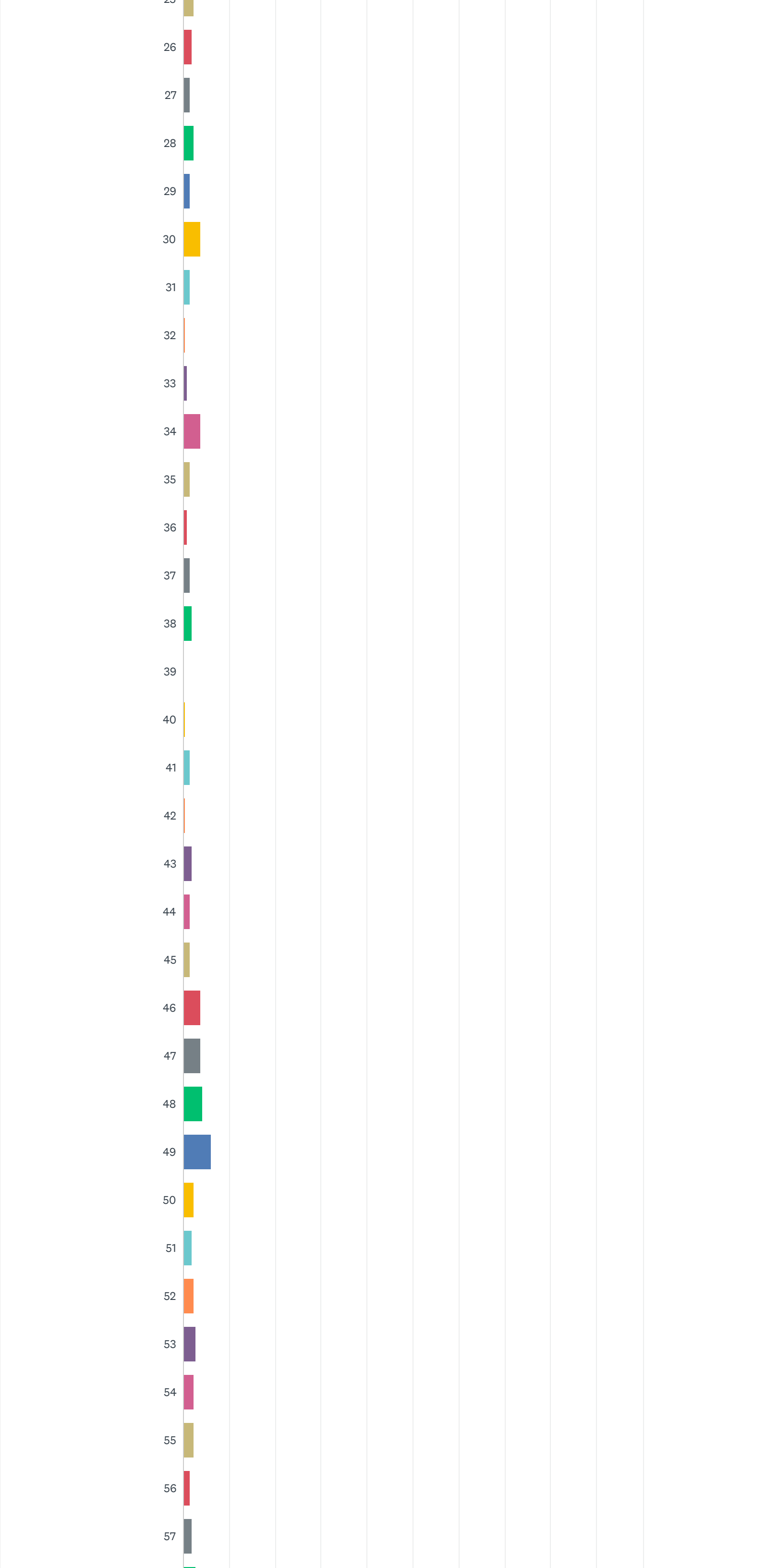

× 

#### Your feedback is important to us

Thank you! We read every response. Privacy notice

Do the analyze visualizations and tools help you gather data insights from your survey?

Yes

No

Continue

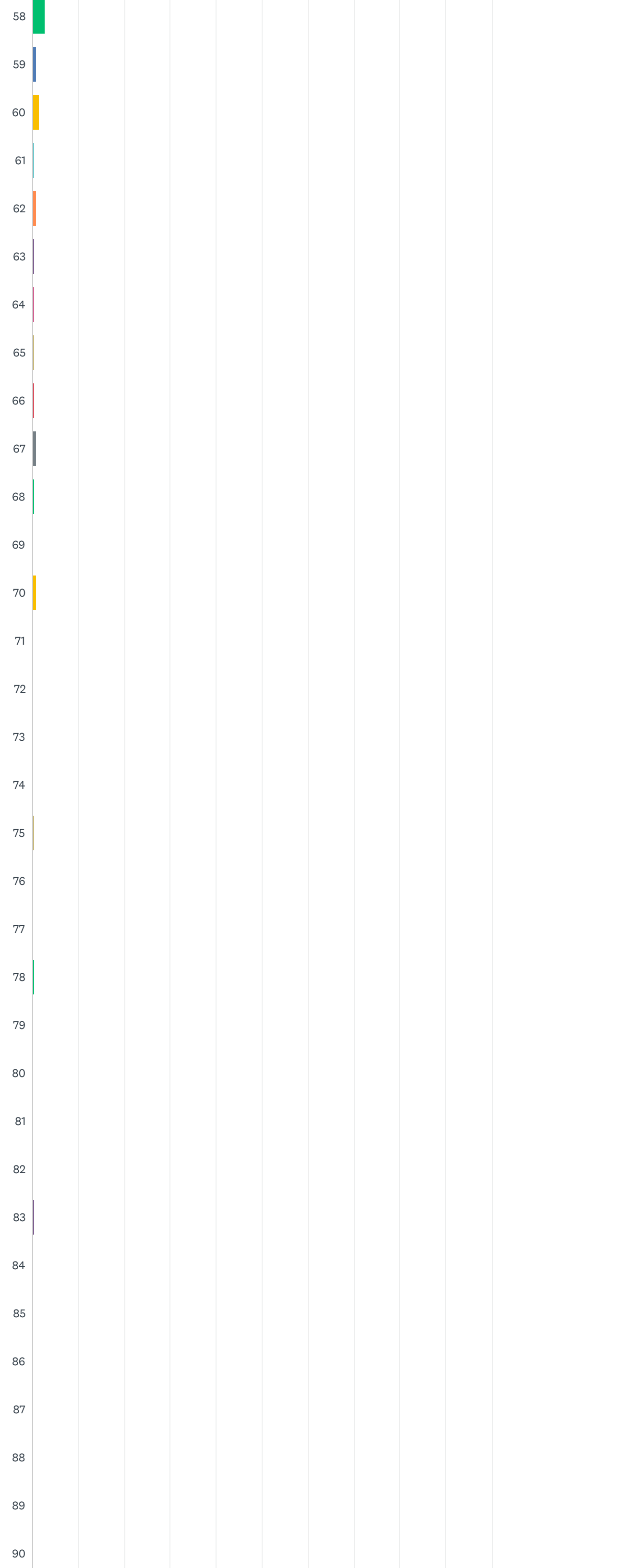

×

#### Your feedback is important to us

Thank you! We read every response. Privacy notice

Do the analyze visualizations and tools help you gather data insights from your survey?

- Yes
- No

Continue

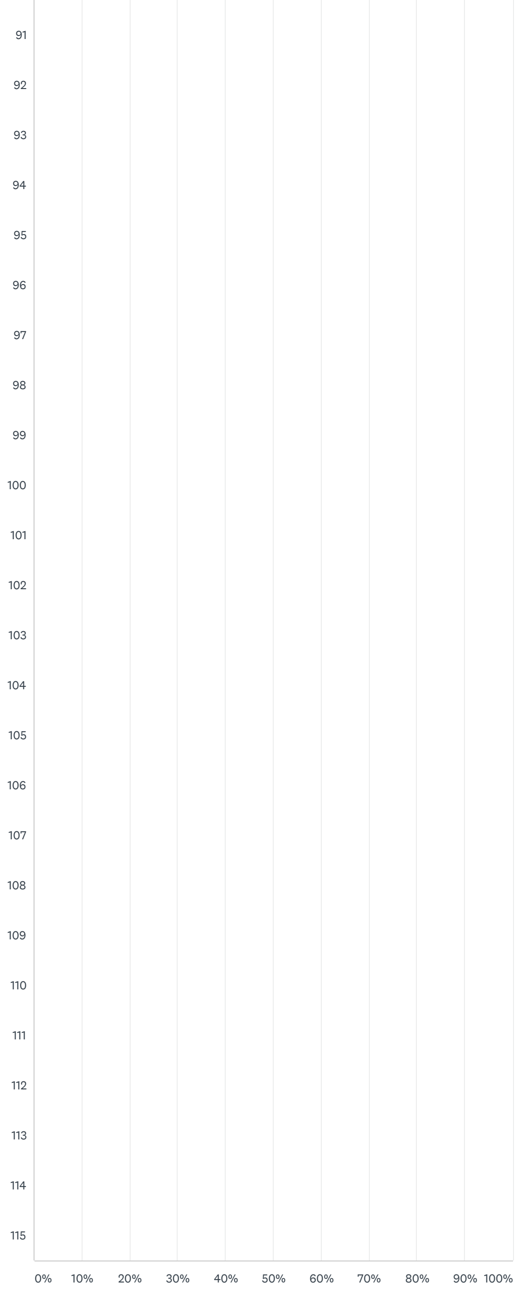


| ANSWER CHOICES | RESPONSES | |
|---|---|---|
| 18 | 5.58% | 12 |
| 19 | 0.93% | 2 |
| 20 | 3.26% | 7 |
| 21 | 4.19% | 9 |
| 22 | 2.79% | 6 |
| 23 | 2.33% | 5 |
| 24 | 1.86% | 4 |
| 25 | 2.33% | 5 |
| 26 | 1.86% | 4 |
| 27 | [illegible] | [illegible] |

ANSWER CHOICES RESPONSES

| | | |
|---|---|---|
| TOTAL | | 215 |
| 27 | 1.40% | 3 |
| 28 | 2.33% | 5 |
| 29 | 1.40% | 3 |
| 30 | 3.72% | 8 |
| 31 | 1.40% | 3 |
| 32 | 0.47% | 1 |
| 33 | 0.93% | 2 |
| 34 | 3.72% | 8 |
| 35 | 1.40% | 3 |
| 36 | 0.93% | 2 |
| 37 | 1.40% | 3 |
| 38 | 1.86% | 4 |
| 39 | 0.00% | 0 |
| 40 | 0.47% | 1 |
| 41 | 1.40% | 3 |
| 42 | 0.47% | 1 |
| 43 | 1.86% | 4 |
| 44 | 1.40% | 3 |
| 45 | 1.40% | 3 |
| 46 | 3.72% | 8 |
| 47 | 3.72% | 8 |
| 48 | 4.19% | 9 |
| 49 | 6.05% | 13 |
| 50 | 2.33% | 5 |
| 51 | 1.86% | 4 |
| 52 | 2.33% | 5 |
| 53 | 2.79% | 6 |
| 54 | 2.33% | 5 |
| 55 | 2.33% | 5 |
| 56 | 1.40% | 3 |
| 57 | 1.86% | 4 |
| 58 | 2.79% | 6 |
| 59 | 0.93% | 2 |
| 60 | 1.40% | 3 |
| 61 | 0.47% | 1 |
| 62 | 0.93% | 2 |
| 63 | 0.47% | 1 |
| 64 | 0.47% | 1 |
| 65 | 0.47% | 1 |
| 66 | 0.47% | 1 |
| 67 | 0.93% | 2 |
| 68 | 0.47% | 1 |
| 69 | 0.00% | 0 |
| 70 | 0.93% | 2 |
| 71 | 0.00% | 0 |
| 72 | 0.00% | 0 |
| 73 | 0.00% | 0 |
| 74 | 0.00% | 0 |
| 75 | 0.47% | 1 |
| 76 | 0.00% | 0 |
| 77 | 0.00% | 0 |
| 78 | 0.47% | 1 |
| 79 | 0.00% | 0 |
| 80 | 0.00% | 0 |
| 81 | 0.00% | 0 |
| 82 | 0.00% | 0 |
| 83 | 0.47% | 1 |
| 84 | 0.00% | 0 |
| ANSWER CHOICES | RESPONSES | |
| [illegible] | 0.00% | 0 |


#### Your feedback is important to us

Thank you! We read every response. Privacy notice

Do the analyze visualizations and tools help you gather data insights from your survey?

Yes

No

Continue

| | | |
|---|---|---|
| 86 | 0.00% | 0 |
| 87 | 0.00% | 0 |
| 88 | 0.00% | 0 |
| 89 | 0.00% | 0 |
| 90 | 0.00% | 0 |
| 91 | 0.00% | 0 |
| 92 | 0.00% | 0 |
| 93 | 0.00% | 0 |
| 94 | 0.00% | 0 |
| 95 | 0.00% | 0 |
| 96 | 0.00% | 0 |
| 97 | 0.00% | 0 |
| 98 | 0.00% | 0 |
| 99 | 0.00% | 0 |
| 100 | 0.00% | 0 |
| 101 | 0.00% | 0 |
| 102 | 0.00% | 0 |
| 103 | 0.00% | 0 |
| 104 | 0.00% | 0 |
| 105 | 0.00% | 0 |
| 106 | 0.00% | 0 |
| 107 | 0.00% | 0 |
| 108 | 0.00% | 0 |
| 109 | 0.00% | 0 |
| 110 | 0.00% | 0 |
| 111 | 0.00% | 0 |
| 112 | 0.00% | 0 |
| 113 | 0.00% | 0 |
| 114 | 0.00% | 0 |
| 115 | 0.00% | 0 |
| TOTAL | | 215 |


×

**Your feedback is important to us**

Thank you! We read every response. Privacy notice

Do the analyze visualizations and tools help you gather data insights from your survey?

- Yes
- No

Continue


Q3 Customize Save as

#### 1. What gender do you identify as?

Answered: 215 Skipped: 796

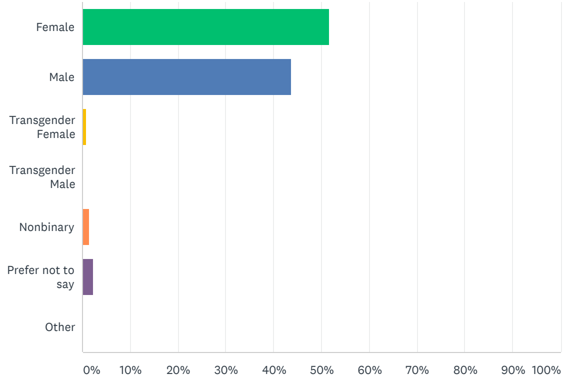


| ANSWER CHOICES | RESPONSES | |
|---|---|---|
| Female | 51.63% | 111 |
| Male | 43.72% | 94 |
| Transgender Female | 0.93% | 2 |

| | | |
|---|---|---|
| Transgender Male | 0.00% | 0 |
| Nonbinary | 1.40% | 3 |
| Prefer not to say | 2.33% | 5 |
| Other | 0.00% | 0 |
| **TOTAL** | | **215** |

Comments (5)


Your feedback is important to us

Thank you! We read every response. Privacy notice

Do the analyze visualizations and tools help you gather data insights from your survey?

Yes

No

Continue


**Q4**

Customize Save as

#### 1. I or my family is ethnically from one or more of the following countries

Answered: 215 Skipped: 796

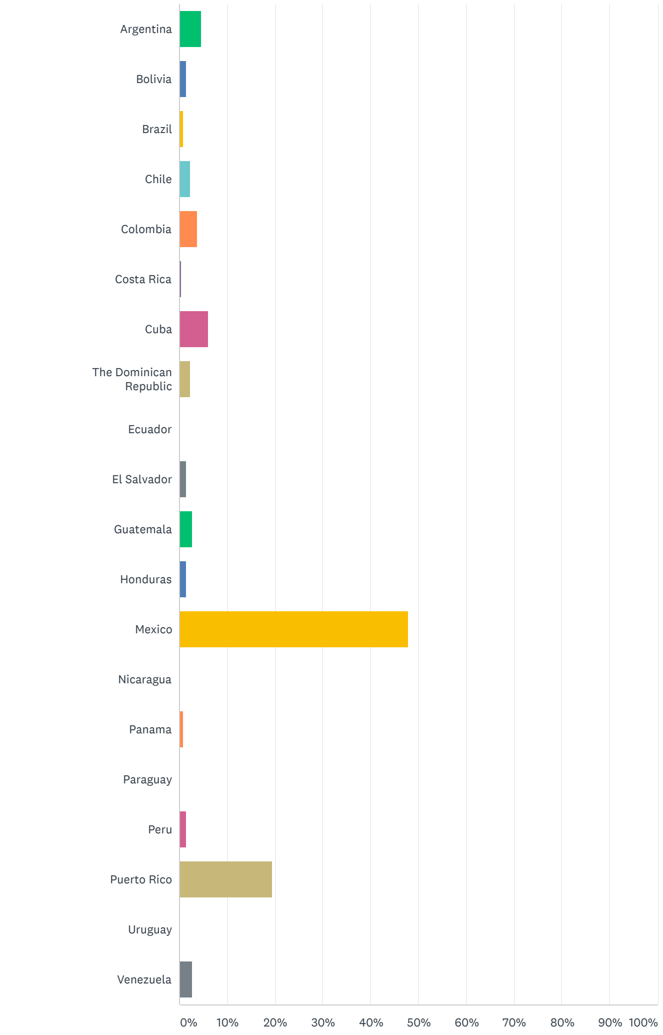


| ANSWER CHOICES | RESPONSES | |
|---|---|---|
| Argentina | 4.65% | 10 |
| Bolivia | 1.40% | 3 |
| Brazil | [illegible] | 2 |

| | | | |
|---|---|---|---|
| ▾ | Chile | 2.33% | 5 |
| ▾ | Colombia | 3.72% | 8 |
| ▾ | Costa Rica | 0.47% | 1 |
| ▾ | Cuba | 6.05% | 13 |
| ▾ | The Dominican Republic | 2.33% | 5 |
| ▾ | Ecuador | 0.00% | 0 |
| ▾ | El Salvador | 1.40% | 3 |
| ▾ | Guatemala | 2.79% | 6 |
| ▾ | Honduras | 1.40% | 3 |
| ▾ | Mexico | 47.91% | 103 |
| ▾ | Nicaragua | 0.00% | 0 |
| ▾ | Panama | 0.93% | 2 |
| ▾ | Paraguay | 0.00% | 0 |
| ▾ | Peru | 1.40% | 3 |
| ▾ | Puerto Rico | 19.53% | 42 |
| ▾ | Uruguay | 0.00% | 0 |
| ▾ | Venezuela | 2.79% | 6 |
| TOTAL | | | 215 |


Your feedback is important to us

Thank you! We read every response. Privacy notice

Do the analyze visualizations and tools help you gather data insights from your survey?

Yes

No

Continue


**Q5** Customize Save as▾

#### 1. What is your yearly income for your household?

Answered: 215 Skipped: 796

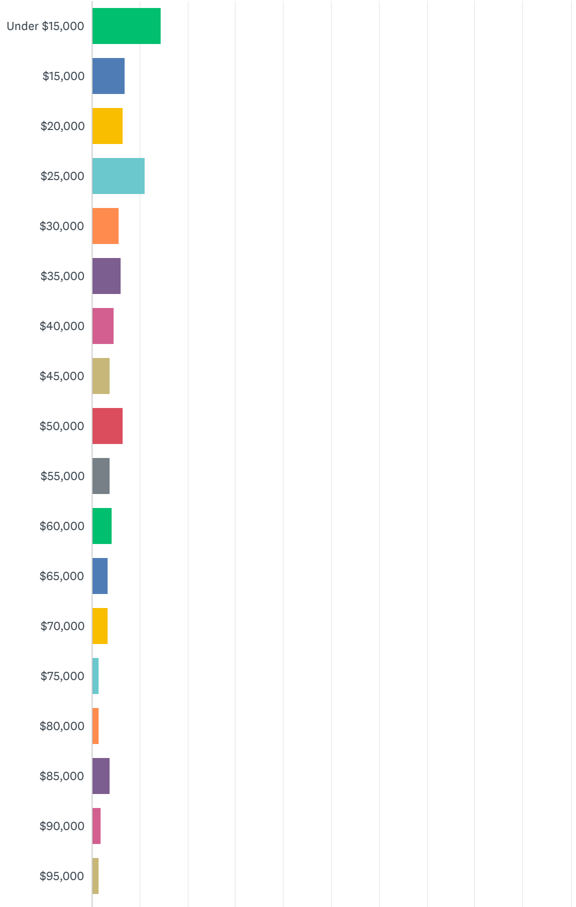

×

Your feedback is important to us

Thank you! We read every response. Privacy notice

Do the analyze visualizations and tools help you gather data insights from your survey?

Yes

No

Continue

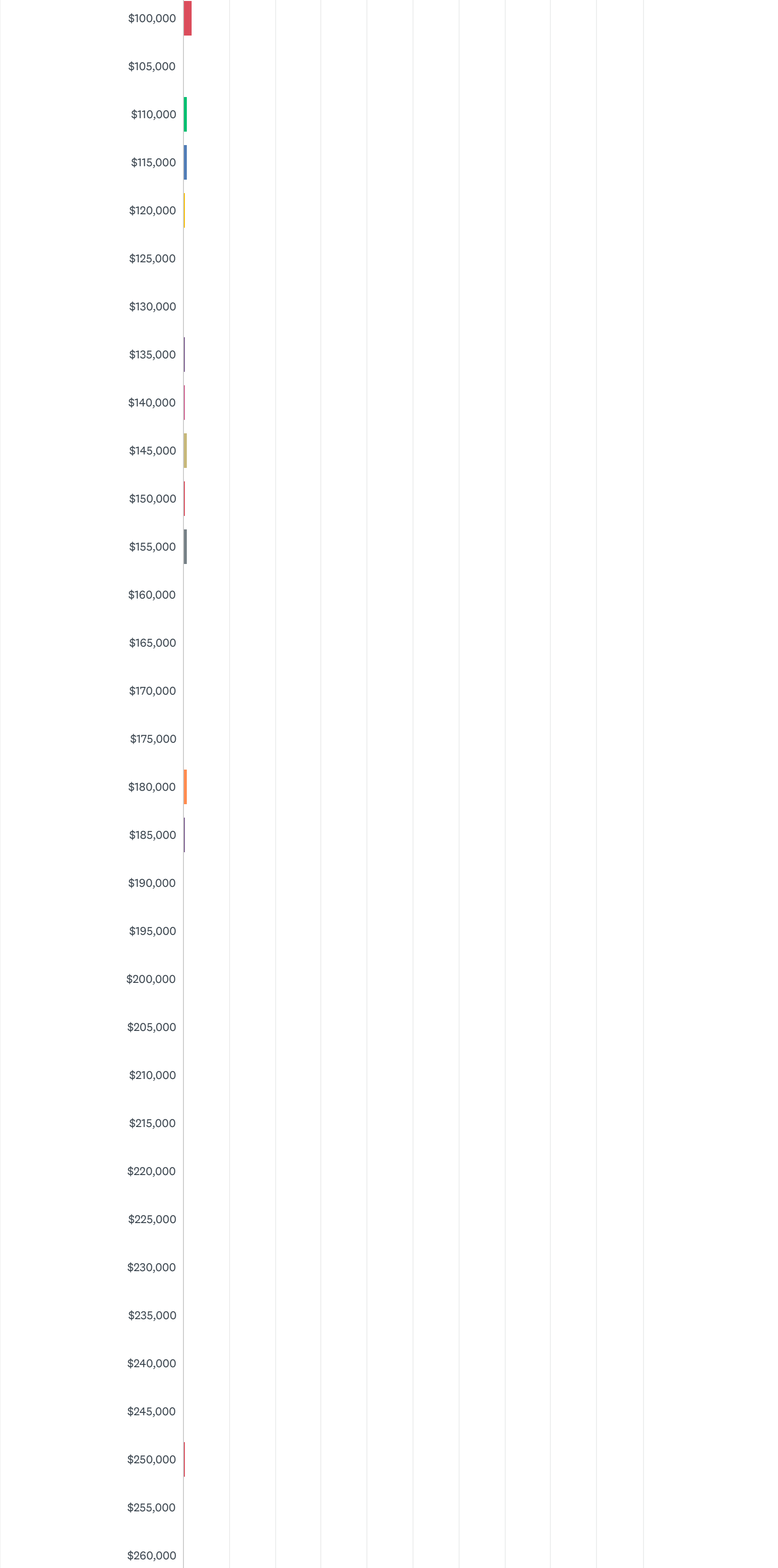

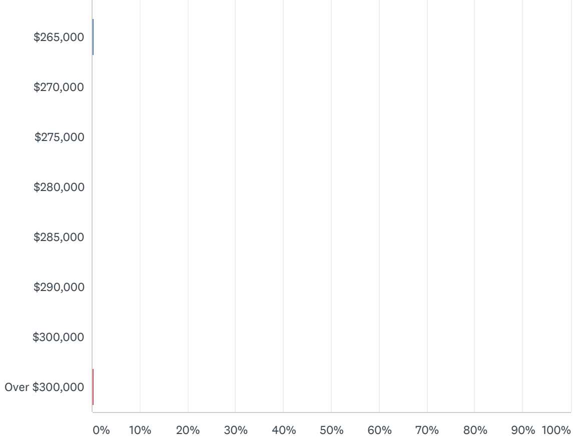


#### Your feedback is important to us

Thank you! We read every response. Privacy notice

Do the analyze visualizations and tools help you gather data insights from your survey?

- Yes
- No

Continue

| ANSWER CHOICES | RESPONSES | |
|---|---|---|
| Under $15,000 | 14.42% | 31 |
| $15,000 | 6.98% | 15 |
| $20,000 | 6.51% | 14 |
| $25,000 | 11.16% | 24 |
| $30,000 | 5.58% | 12 |
| $35,000 | 6.05% | 13 |
| $40,000 | 4.65% | 10 |
| $45,000 | 3.72% | 8 |
| $50,000 | 6.51% | 14 |
| $55,000 | 3.72% | 8 |
| $60,000 | 4.19% | 9 |
| $65,000 | 3.26% | 7 |
| $70,000 | 3.26% | 7 |
| $75,000 | 1.40% | 3 |
| $80,000 | 1.40% | 3 |
| $85,000 | 3.72% | 8 |
| $90,000 | 1.86% | 4 |
| $95,000 | 1.40% | 3 |
| $100,000 | 1.86% | 4 |
| $105,000 | 0.00% | 0 |
| $110,000 | 0.93% | 2 |
| $115,000 | 0.93% | 2 |
| $120,000 | 0.47% | 1 |
| $125,000 | 0.00% | 0 |
| $130,000 | 0.00% | 0 |
| $135,000 | 0.47% | 1 |
| $140,000 | 0.47% | 1 |
| $145,000 | 0.93% | 2 |
| $150,000 | 0.47% | 1 |
| $155,000 | 0.93% | 2 |
| $160,000 | 0.00% | 0 |
| $165,000 | 0.00% | 0 |
| $170,000 | 0.00% | 0 |
| $175,000 | 0.00% | 0 |
| $180,000 | 0.93% | 2 |
| $185,000 | 0.47% | 1 |
| $190,000 | 0.00% | 0 |
| $195,000 | 0.00% | 0 |
| $200,000 | 0.00% | 0 |
| $205,000 | 0.00% | 0 |

ANSWER CHOICES RESPONSES

| | | |
|---|---|---|
| $210,000 | 0.00% | 0 |
| $215,000 | 0.00% | 0 |
| $220,000 | 0.00% | 0 |
| $225,000 | 0.00% | 0 |
| $230,000 | 0.00% | 0 |
| $235,000 | 0.00% | 0 |
| $240,000 | 0.00% | 0 |
| $245,000 | 0.00% | 0 |
| $250,000 | 0.47% | 1 |
| $255,000 | 0.00% | 0 |
| $260,000 | 0.00% | 0 |
| $265,000 | 0.47% | 1 |
| $270,000 | 0.00% | 0 |
| $275,000 | 0.00% | 0 |
| $280,000 | 0.00% | 0 |
| $285,000 | 0.00% | 0 |
| $290,000 | 0.00% | 0 |
| $300,000 | 0.00% | 0 |
| Over $300,000 | 0.47% | 1 |
| TOTAL | | 215 |


×

**Your feedback is important to us**

Thank you! We read every response. Privacy notice

Do the analyze visualizations and tools help you gather data insights from your survey?

Yes

No

Continue


## Q6

Customize Save as

### What is the highest level of education you have completed?

Answered: 215 Skipped: 796

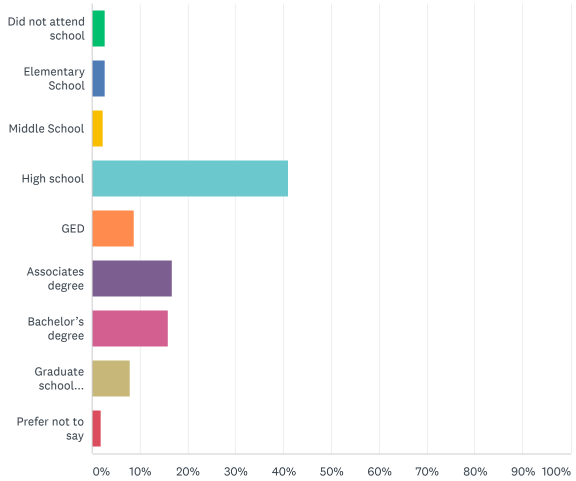


| ANSWER CHOICES | RESPONSES | |
|---|---|---|
| Did not attend school | 2.79% | 6 |
| Elementary School | 2.79% | 6 |
| Middle School | 2.33% | 5 |
| High school | 40.93% | 88 |
| GED | 8.84% | 19 |
| Associates degree | 16.74% | 36 |
| Bachelor’s degree | 15.81% | 34 |
| Graduate school (Master’s Degree, MBA, JD, PhD, MD, DO, DDS, etc.) | 7.91% | 17 |
| Prefer not to say | 1.86% | 4 |
| TOTAL | | 215 |

Your feedback is important to us
Thank you! We read every response. Privacy notice
Do the analyze visualizations and tools help you gather data insights from your survey?
Yes
No
Continue

Q7

Customize Save as

#### What is your political affiliation?

Answered: 215 Skipped: 796

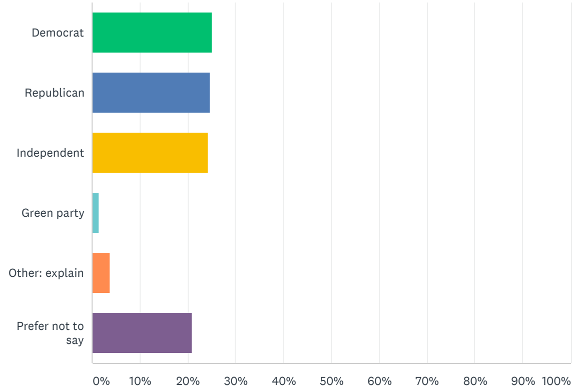


| ANSWER CHOICES | RESPONSES | |
|---|---|---|
| Democrat | 25.12% | 54 |
| Republican | 24.65% | 53 |
| Independent | 24.19% | 52 |
| Green party | 1.40% | 3 |
| Other: explain | 3.72% | 8 |
| Prefer not to say | 20.93% | 45 |
| TOTAL | | 215 |

Comments (10)

Page 3

Q8

Customize Save as

#### What are your top 3 reasons for not receiving the COVID-19 vaccine?

Answered: 211 Skipped: 800

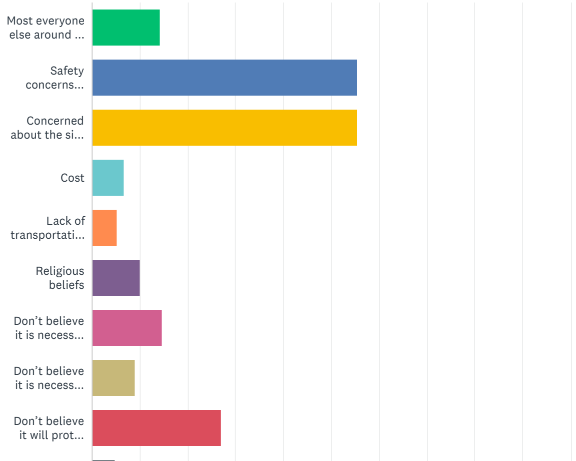

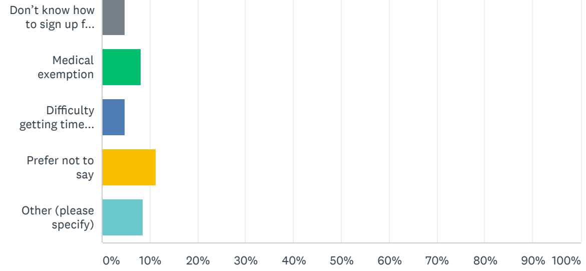


| ANSWER CHOICES | | RESPONSES | |
|---|---|---|---|
| Most everyone else around me already received the vaccine | | 14.22% | 30 |
| Safety concerns regarding the vaccine contents | | 55.45% | 117 |
| Concerned about the side effects | | 55.45% | 117 |
| Cost | | 6.64% | 14 |
| Lack of transportation to vaccination site | | 5.21% | 11 |
| Religious beliefs | | 9.95% | 21 |
| Don't believe it is necessary (previously diagnosed with COVID-19 by a healthcare provider) | | 14.69% | 31 |
| Don't believe it is necessary (suspect I previously had COVID-19) | | 9.00% | 19 |
| Don't believe it will protect me from COVID-19 | | 27.01% | 57 |
| Don't know how to sign up for a vaccination | | 4.74% | 10 |
| Medical exemption | | 8.06% | 17 |
| Difficulty getting time off from work | | 4.74% | 10 |
| Prefer not to say | | 11.37% | 24 |
| Other (please specify) | Responses | 8.53% | 18 |
| **Total Respondents: 211** | | | |


×

Your feedback is important to us

Thank you! We read every response. Privacy notice

Do the analyze visualizations and tools help you gather data insights from your survey?

Yes

No

Continue


Q9

Customize Save as

#### How likely would you be to take the COVID vaccine if it were a pill?

Answered: 212 Skipped: 799

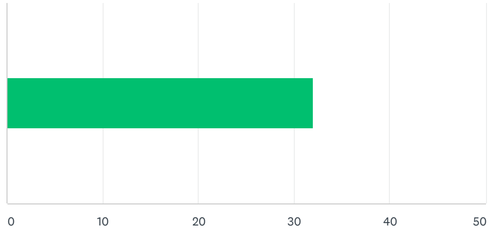


| ANSWER CHOICES | AVERAGE NUMBER | TOTAL NUMBER | RESPONSES |
|---|---|---|---|
| Responses | 32 | 6,785 | 212 |
| **Total Respondents: 212** | | | |

Q10

Customize Save as

#### How many of the 30 people you interact with (non-virtually) the most each week (your bubble) have received the COVID vaccine?

Answered: 212 Skipped: 799

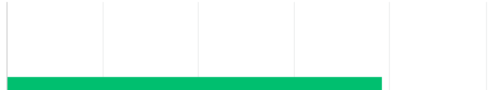

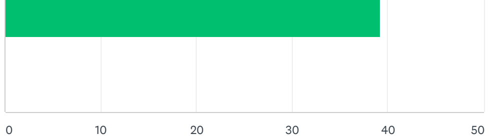


| ANSWER CHOICES | AVERAGE NUMBER | TOTAL NUMBER | RESPONSES |
|---|---|---|---|
| Responses | 39 | 8,316 | 212 |

RESPONSES (212) TAGS (0)

Add tags Filter by tag Search responses

Showing **212** responses

39
8/1/2021 2:51 PM View respondent's answers Add tags

82
8/1/2021 12:37 PM View respondent's answers Add tags

35
8/1/2021 12:29 PM View respondent's answers Add tags

0
8/1/2021 11:51 AM View respondent's answers Add tags

Total Respondents: 212


#### Your feedback is important to us

Thank you! We read every response. Privacy notice

Do the analyze visualizations and tools help you gather data insights from your survey?

Yes

No

Continue


Q11 Customize Save as

#### How likely would you be to receive the vaccine if the majority of your bubble received the vaccine?

Answered: 212 Skipped: 799

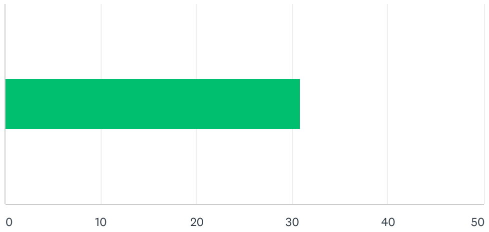


| ANSWER CHOICES | AVERAGE NUMBER | TOTAL NUMBER | RESPONSES |
|---|---|---|---|
| Responses | 31 | 6,548 | 212 |

Total Respondents: 212

Q12 Customize Save as

#### Within the past month, how often have you attended in person or virtual religious services?

Answered: 212 Skipped: 799

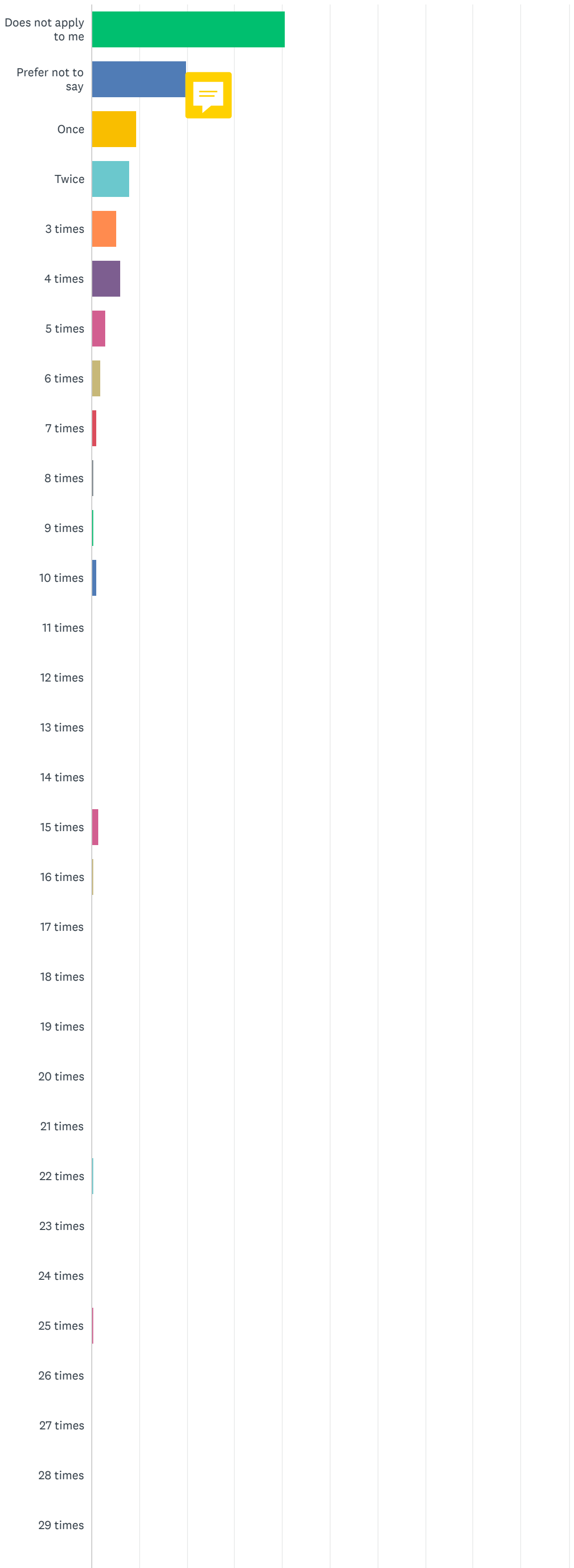


×

**Your feedback is important to us**

Thank you! We read every response. Privacy notice

Do the analyze visualizations and tools help you gather data insights from your survey?

- Yes
- No

Continue

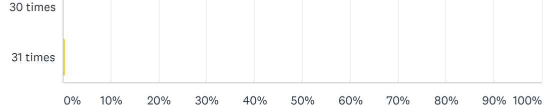


| ANSWER CHOICES | RESPONSES | |
|---|---|---|
| Does not apply to me | 40.57% | 86 |
| Prefer not to say | 19.81% | 42 |
| Once | 9.43% | 20 |
| Twice | 8.02% | 17 |
| 3 times | 5.19% | 11 |
| 4 times | 6.13% | 13 |
| 5 times | 2.83% | 6 |
| 6 times | 1.89% | 4 |
| 7 times | 0.94% | 2 |
| 8 times | 0.47% | 1 |
| 9 times | 0.47% | 1 |
| 10 times | 0.94% | 2 |
| 11 times | 0.00% | 0 |
| 12 times | 0.00% | 0 |
| 13 times | 0.00% | 0 |
| 14 times | 0.00% | 0 |
| 15 times | 1.42% | 3 |
| 16 times | 0.47% | 1 |
| 17 times | 0.00% | 0 |
| 18 times | 0.00% | 0 |
| 19 times | 0.00% | 0 |
| 20 times | 0.00% | 0 |
| 21 times | 0.00% | 0 |
| 22 times | 0.47% | 1 |
| 23 times | 0.00% | 0 |
| 24 times | 0.00% | 0 |
| 25 times | 0.47% | 1 |
| 26 times | 0.00% | 0 |
| 27 times | 0.00% | 0 |
| 28 times | 0.00% | 0 |
| 29 times | 0.00% | 0 |
| 30 times | 0.00% | 0 |
| 31 times | 0.47% | 1 |
| TOTAL | | 212 |

×

Your feedback is important to us

Thank you! We read every response. Privacy notice

Do the analyze visualizations and tools help you gather data insights from your survey?

Yes

No

Continue

Page 4

**Q13**

Customize Save as

#### The COVID-19 vaccine will make me infertile.

Answered: 209 Skipped: 802

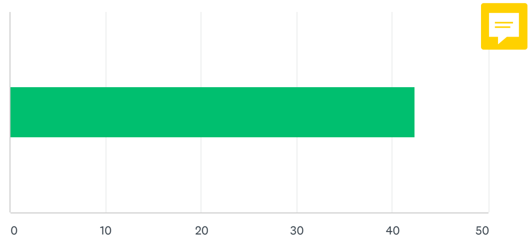

| ANSWER CHOICES | AVERAGE NUMBER | TOTAL NUMBER | RESPONSES |
|---|---|---|---|
| Responses | 42 | 8,850 | 209 |
| Total Respondents: 209 | | | |

Your feedback is important to us
Thank you! We read every response. Privacy notice
Do the analyze visualizations and tools help you gather data insights from your survey?
Yes
No
Continue

Q14

Customize Save as

#### The developers of the COVID-19 vaccine rushed the development and cut corners.

Answered: 209 Skipped: 802

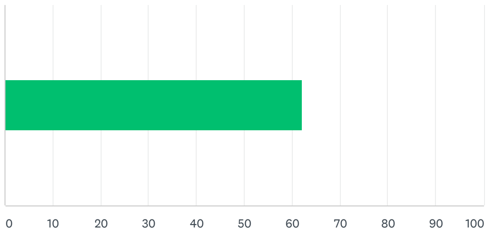


| ANSWER CHOICES | AVERAGE NUMBER | TOTAL NUMBER | RESPONSES |
|---|---|---|---|
| Responses | 62 | 13,004 | 209 |
| Total Respondents: 209 | | | |

Q15

Customize Save as

#### The COVID-19 vaccine is just the virus and will infect you with the disease.

Answered: 209 Skipped: 802

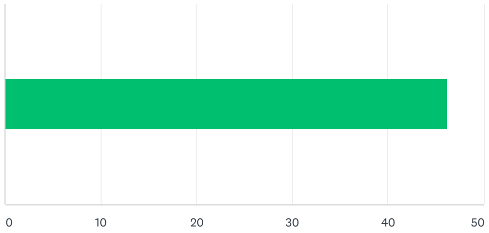


| ANSWER CHOICES | AVERAGE NUMBER | TOTAL NUMBER | RESPONSES |
|---|---|---|---|
| Responses | 46 | 9,653 | 209 |
| Total Respondents: 209 | | | |

Q16

Customize Save as

#### If I’ve already had COVID-19, I don’t need the vaccine.

Answered: 209 Skipped: 802

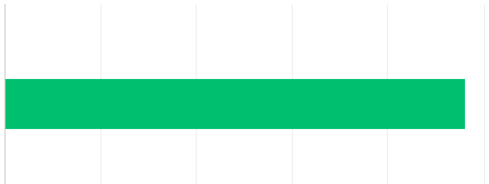

| ANSWER CHOICES | AVERAGE NUMBER | TOTAL NUMBER | RESPONSES |
|---|---|---|---|
| Responses | 48 | 10,060 | 209 |
| Total Respondents: 209 | | | |

×

Your feedback is important to us

Thank you! We read every response. Privacy notice

Do the analyze visualizations and tools help you gather data insights from your survey?

Yes

No

Continue

Q17

Customize Save as

#### The COVID-19 vaccine does not work.

Answered: 209 Skipped: 802

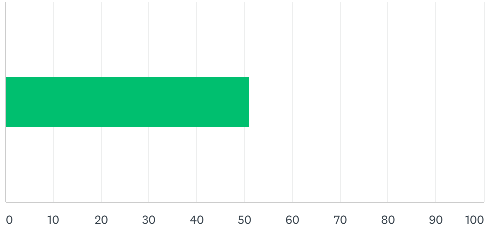


| ANSWER CHOICES | AVERAGE NUMBER | TOTAL NUMBER | RESPONSES |
|---|---|---|---|
| Responses | 51 | 10,667 | 209 |
| Total Respondents: 209 | | | |

Q18

Customize Save as

#### COVID-19 is a myth.

Answered: 209 Skipped: 802

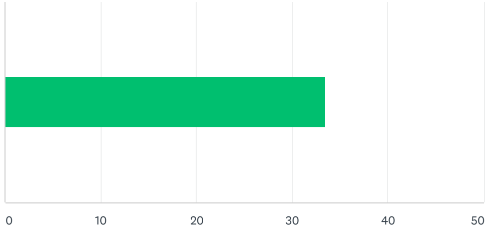


| ANSWER CHOICES | AVERAGE NUMBER | TOTAL NUMBER | RESPONSES |
|---|---|---|---|
| Responses | 33 | 6,985 | 209 |
| Total Respondents: 209 | | | |

Q19

Customize Save as

#### The vaccine isn't necessary because COVID-19 has a low mortality rate.

Answered: 209 Skipped: 802

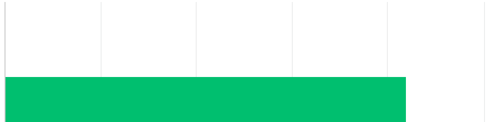

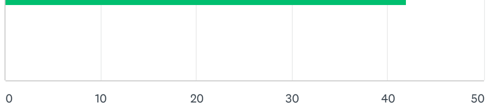


| ANSWER CHOICES | AVERAGE NUMBER | TOTAL NUMBER | RESPONSES |
|---|---|---|---|
| Responses | 42 | 8,763 | 209 |
| Total Respondents: 209 | | | |


×

Your feedback is important to us

Thank you! We read every response. Privacy notice

Do the analyze visualizations and tools help you gather data insights from your survey?

Yes

No

Continue


Q20

Customize Save as

#### I don’t need the vaccine because everyone else around me has already received it.

Answered: 209 Skipped: 802

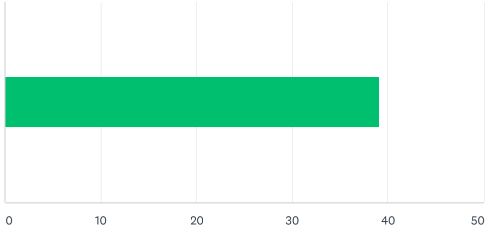


| ANSWER CHOICES | AVERAGE NUMBER | TOTAL NUMBER | RESPONSES |
|---|---|---|---|
| Responses | 39 | 8,166 | 209 |
| Total Respondents: 209 | | | |

Q21

Customize Save as

#### I only need the vaccine if I want to travel out of the country

Answered: 209 Skipped: 802

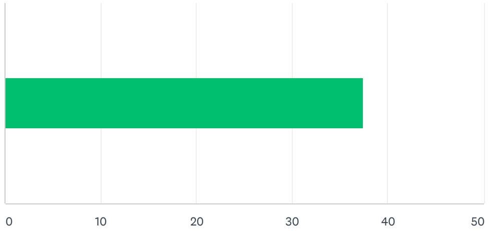


| ANSWER CHOICES | AVERAGE NUMBER | TOTAL NUMBER | RESPONSES |
|---|---|---|---|
| Responses | 37 | 7,819 | 209 |
| Total Respondents: 209 | | | |

Q22

Customize Save as

#### The COVID-19 vaccine will change parts of my DNA.

Answered: 209 Skipped: 802

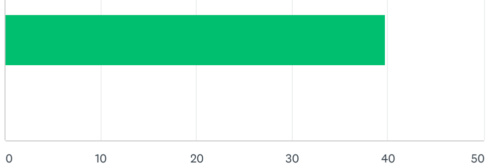


| ANSWER CHOICES | AVERAGE NUMBER | TOTAL NUMBER | RESPONSES |
|---|---|---|---|
| Responses | 40 | 8,308 | 209 |
| Total Respondents: 209 | | | |

×

#### Your feedback is important to us

Thank you! We read every response. Privacy notice

Do the analyze visualizations and tools help you gather data insights from your survey?

Yes

No

Continue

Q23

Customize Save as

#### For what ages and groups do the COVID-19 vaccine benefits exceed the risks or side effects? Check all that apply.

Answered: 209 Skipped: 802

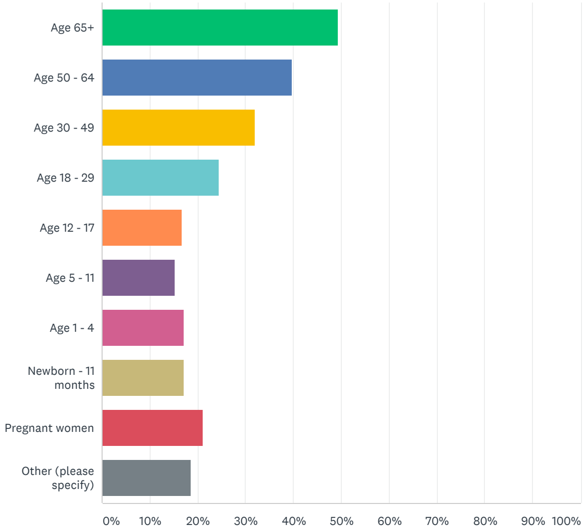


| ANSWER CHOICES | | RESPONSES | |
|---|---|---|---|
| Age 65+ | | 49.28% | 103 |
| Age 50 - 64 | | 39.71% | 83 |
| Age 30 - 49 | | 32.06% | 67 |
| Age 18 - 29 | | 24.40% | 51 |
| Age 12 - 17 | | 16.75% | 35 |
| Age 5 - 11 | | 15.31% | 32 |
| Age 1 - 4 | | 17.22% | 36 |
| Newborn - 11 months | | 17.22% | 36 |
| Pregnant women | | 21.05% | 44 |
| Other (please specify) | Responses | 18.66% | 39 |
| Total Respondents: 209 | | | |

Q24

Customize Save as

#### Which of the following are your top 3 primary sources of news?

#### Which of the following are your top 3 primary sources of news?

Answered: 208 Skipped: 803

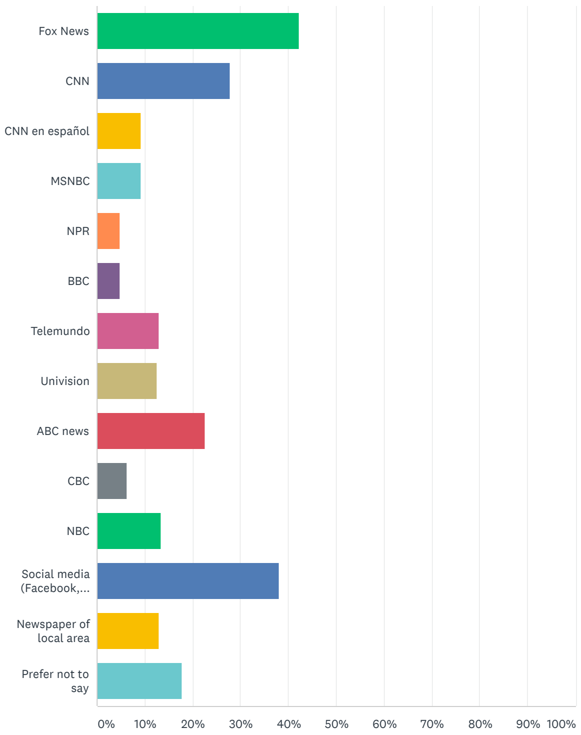


| ANSWER CHOICES | RESPONSES | |
|---|---|---|
| Fox News | 42.31% | 88 |
| CNN | 27.88% | 58 |
| CNN en español | 9.13% | 19 |
| MSNBC | 9.13% | 19 |
| NPR | 4.81% | 10 |
| BBC | 4.81% | 10 |
| Telemundo | 12.98% | 27 |
| Univision | 12.50% | 26 |
| ABC news | 22.60% | 47 |
| CBC | 6.25% | 13 |
| NBC | 13.46% | 28 |
| Social media (Facebook, Twitter, Instagram, etc.) | 37.98% | 79 |
| Newspaper of local area | 12.98% | 27 |
| Prefer not to say | 17.79% | 37 |
| Total Respondents: 208 | | |


×

Your feedback is important to us

Thank you! We read every response. Privacy notice

Do the analyze visualizations and tools help you gather data insights from your survey?

Yes

No

Continue


### Q25

Customize Save as

#### If the election were held today, how likely would you be to vote for Donald Trump or Joe Biden?

Answered: 209 Skipped: 802

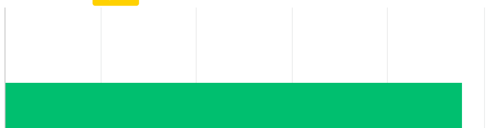

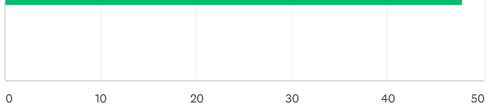


| ANSWER CHOICES | AVERAGE NUMBER | TOTAL NUMBER | RESPONSES |
|---|---|---|---|
| Responses | 48 | 9,980 | 209 |
| Total Respondents: 209 | | | |


×

**Your feedback is important to us**

Thank you! We read every response. Privacy notice

Do the analyze visualizations and tools help you gather data insights from your survey?

- Yes
- No

Continue


Page 5: SurveyMonkey Audience

**Q26**

Customize  Save as

#### Device Type

Answered: 980    Skipped: 31

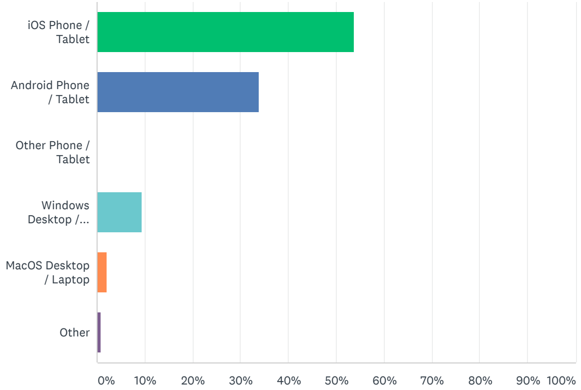


| ANSWER CHOICES | RESPONSES | |
|---|---|---|
| iOS Phone / Tablet | 53.67% | 526 |
| Android Phone / Tablet | 33.88% | 332 |
| Other Phone / Tablet | 0.00% | 0 |
| Windows Desktop / Laptop | 9.39% | 92 |
| MacOS Desktop / Laptop | 2.14% | 21 |
| Other | 0.92% | 9 |
| TOTAL | | 980 |

**Q27**

Customize  Save as

#### Age

Answered: 980    Skipped: 31

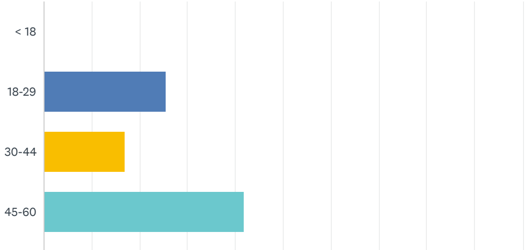

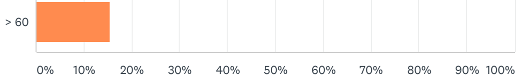


| ANSWER CHOICES | RESPONSES | |
|---|---|---|
| < 18 | 0.00% | 0 |
| 18-29 | 25.61% | 251 |
| 30-44 | 17.04% | 167 |
| 45-60 | 41.94% | 411 |
| > 60 | 15.41% | 151 |
| TOTAL | | 980 |


×

Your feedback is important to us

Thank you! We read every response. Privacy notice

Do the analyze visualizations and tools help you gather data insights from your survey?

Yes

No

Continue


Q28

Customize Save as

#### Region

Answered: 962 Skipped: 49

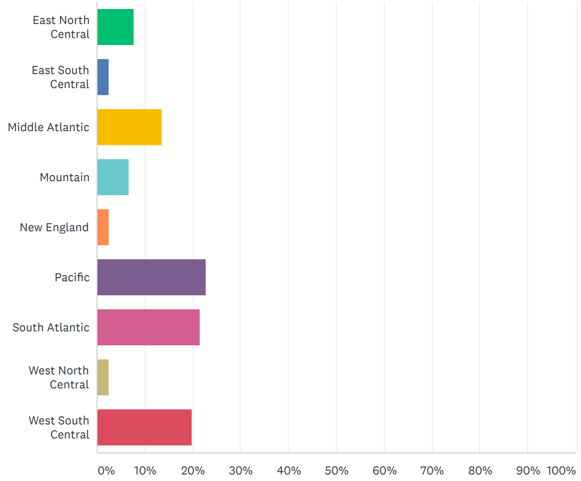


| ANSWER CHOICES | RESPONSES | |
|---|---|---|
| East North Central | 7.80% | 75 |
| East South Central | 2.49% | 24 |
| Middle Atlantic | 13.51% | 130 |
| Mountain | 6.65% | 64 |
| New England | 2.49% | 24 |
| Pacific | 22.87% | 220 |
| South Atlantic | 21.62% | 208 |
| West North Central | 2.60% | 25 |
| West South Central | 19.96% | 192 |
| TOTAL | | 962 |

Q29

Customize Save as

#### Gender

Answered: 980 Skipped: 31

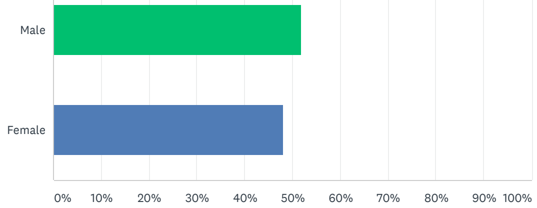


| ANSWER CHOICES | RESPONSES | |
|---|---|---|
| Male | 51.94% | 509 |
| Female | 48.06% | 471 |
| TOTAL | | 980 |


×

Your feedback is important to us

Thank you! We read every response. Privacy notice

Do the analyze visualizations and tools help you gather data insights from your survey?

Yes

No

Continue


Q30

Customize Save as

#### Household Income

Answered: 980 Skipped: 31

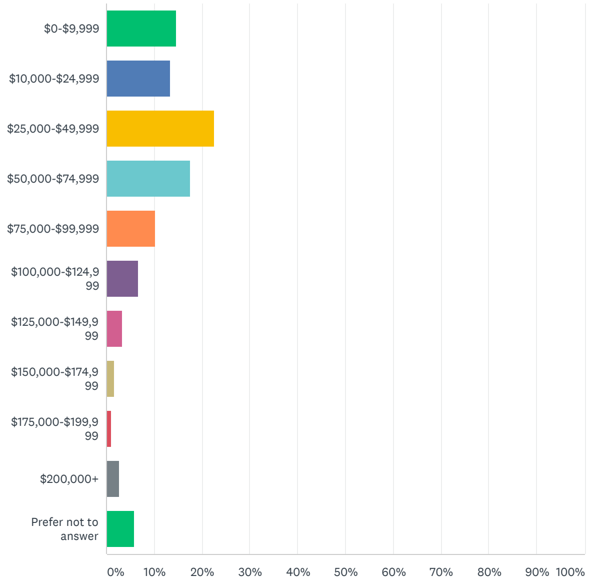


| ANSWER CHOICES | RESPONSES | |
|---|---|---|
| $0-$9,999 | 14.69% | 144 |
| $10,000-$24,999 | 13.37% | 131 |
| $25,000-$49,999 | 22.55% | 221 |
| $50,000-$74,999 | 17.65% | 173 |
| $75,000-$99,999 | 10.20% | 100 |
| $100,000-$124,999 | 6.63% | 65 |
| $125,000-$149,999 | 3.37% | 33 |
| $150,000-$174,999 | 1.73% | 17 |
| $175,000-$199,999 | 1.12% | 11 |
| $200,000+ | 2.76% | 27 |
| Prefer not to answer | 5.92% | 58 |
| TOTAL | | 980 |

ENGLISH

About SurveyMonkey • Careers • Developers • Privacy Notice • California Privacy Notice • Email Opt-In • Help • Cookies Notice • Do Not Sell My Personal Information
Copyright © 1999-2021 SurveyMonkey

Your feedback is important to us
Thank you! We read every response. Privacy notice

Do the analyze visualizations and tools help you gather data insights from your survey?

Yes
No

Continue
